# Associations of schizophrenia with arrhythmic disorders and electrocardiogram traits: an in-depth genetic exploration of population samples

**DOI:** 10.1101/2023.05.21.23290286

**Authors:** Jorien L Treur, Anaïs B Thijssen, Dirk JA Smit, Rafik Tadros, Rada R Veeneman, Damiaan Denys, Jentien M Vermeulen, Julien Barc, Jacob Bergstedt, Joëlle A Pasman, Connie R Bezzina, Karin J H Verweij

## Abstract

**Background:** An important contributor to the decreased life expectancy of individuals with schizophrenia is sudden cardiac death. While arrhythmic disorders play an important role in this, the nature of the relation between schizophrenia and arrhythmia is not fully understood.

**Methods:** We leveraged summary-level data of large-scale genome-wide association studies of schizophrenia (53,386 cases 77,258 controls), arrhythmic disorders (atrial fibrillation, 55,114 cases 482,295 controls; Brugada syndrome, 2,820 cases 10,001 controls) and electrocardiogram traits (heart rate (variability), PR interval, QT interval, JT interval, and QRS duration, n=46,952-293,051). First, we examined shared genetic liability by assessing global and local genetic correlations and conducting functional annotation. Next, we explored bidirectional causal relations between schizophrenia and arrhythmic disorders and electrocardiogram traits using Mendelian randomization.

**Outcomes:** There was no evidence for global genetic correlations, except between schizophrenia and Brugada (r_g_=0·14, *p=*4·0E-04). In contrast, strong positive and negative local genetic correlations between schizophrenia and all cardiac traits were found across the genome. In the strongest associated regions, genes related to immune system and viral response mechanisms were overrepresented. Mendelian randomization indicated a causal, increasing effect of liability to schizophrenia on Brugada syndrome (OR=1·15, *p=*0·009) and heart rate during activity (beta=0·25, *p=*0·015).

**Interpretation:** While there was little evidence for global genetic correlations, specific genomic regions and biological pathways important for both schizophrenia and arrhythmic disorders and electrocardiogram traits emerged. The putative causal effect of liability to schizophrenia on Brugada warrants increased cardiac monitoring and potentially early medical intervention in patients with schizophrenia.

**Funding:** European Research Council Starting Grant.

## Background

Individuals with a serious mental illness have a markedly shorter life expectancy than individuals from the general population. This life expectancy gap is especially stark for patients with schizophrenia, who are expected to live, on average, 10–20 years shorter than individuals without mental illness.^1,2^ While some of these life years lost can be attributed to manifestations of the psychological symptoms, such as suicide^3^, another important cause of premature death is cardiovascular disease.^4^ The risk of sudden cardiac death is ∼10 times higher in individuals with schizophrenia-spectrum disorders compared to the general population.^5,6^ Sudden cardiac death can be the result of structural disorders such as coronary artery disease, but arrhythmic disorders (electrophysiological abnormalities) also play an important role. Patients with schizophrenia show increased rates of arrhythmia, as measured with an electrocardiogram (ECG).^5,7–10^ The most common arrhythmic disorder is atrial fibrillation which, over time, can lead to remodelling of the heart’s ventricles and thereby make it more susceptible to ventricular fibrillation and sudden cardiac death.^11,12^ Brugada syndrome, a rare arrhythmia with a population prevalence of 0·05%^13^, is also more common among individuals with schizophrenia.^7,14^ It is characterized by ST-segment elevation in ECG recordings and associated with an increased risk of sudden death in young adulthood.^15^ While anti-psychotic medication can have cardiac side-effects, its use does not explain these associations. Patients with schizophrenia who do not use medication and young people with a first episode of psychosis also show increased arrhythmia rates.^7,16–18^ Currently, it is poorly understood why schizophrenia is associated with arrhythmia, with epidemiological and clinical studies being hampered by the low prevalence of the variables of interest.

A potential mechanism is shared genetic risk factors, such that genetic variants that convey a higher risk of developing schizophrenia also increase the risk of arrhythmia. In the most recent genome-wide association study (GWAS) of schizophrenia, some of the strongest associations were found with single nucleotide polymorphisms (SNPs) that lie in genes coding for ion channels (mainly voltage-gated calcium channels).^19^ Interestingly, these are also involved in cardiac electrical function and the development of arrhythmia.^20^ To formally assess shared genetic risk, a genetic correlation can be computed, which estimates the overlap between genetic variants that are involved in susceptibility to two traits.^21^ One study found no evidence for genetic correlation between schizophrenia and a range of cardiovascular outcomes (including blood pressure, coronary artery disease, heart rate variability, and heart failure^22^), while another found modest correlations between schizophrenia and cardio-metabolic traits (including lipid levels, BMI and coronary artery disease) but only when selecting lower-frequency genetic variants.^23^ This lack of evidence for (strong) genetic correlation is striking, given that schizophrenia and cardiovascular disease are strongly correlated phenotypically. One explanation may be that genetic correlation only occurs in specific regions of the genome or in opposing directions. This would not be picked up with a global correlation as this measure aggregates associations across the entire genome into a single measure. Sophisticated methods to assess *local* genetic correlations^24^ and the function of shared biological pathways^25^ are available, but have scarcely been applied.

Another potential mechanism for why schizophrenia is associated with arrhythmia is that there are *causal* effects. The most intuitive direction of causality is that schizophrenia increases arrhythmia risk, potentially due to the systemic effects that schizophrenia has on the body and the autonomic nervous system (which also controls the heart’s electrophysiological function).^22,26^ Reverse causal effects are also possible. A longitudinal study in >1 million men showed that a higher heart rate in adolescence increased risk of developing psychosis in adulthood.^27^ High heart rate could represent an early marker of psychotic disorder, but the authors speculated that it could also be a causal risk factor.^28^ Mendelian randomization (MR) mimics a randomized controlled trial (which is not feasible here) by using specific genetic variants as instrumental variables, or ‘proxies’, to test causal effects of a proposed risk factor (‘exposure’) on an outcome. With MR, a subset of significant genetic variants which are strongly and robustly predictive of the exposure is selected. Because genetic variants are randomly passed on from parents to offspring, bias from confounders can be (largely) circumvented. Using MR, we recently found evidence for a causal effect of liability to schizophrenia on heart failure.^22^ The latest availability of large GWASs on arrhythmic disorders and ECG traits now makes it possible to comprehensively assess the causal relation of schizophrenia with arrhythmia.^29–34^

In this pre-registered study (https://osf.io/fe4ms), we assess shared genetic risk factors of schizophrenia with arrhythmic disorders and ECG traits as well as specific biological pathways responsible for such shared liability, and explore potential causal effects of schizophrenia with arrhythmic disorders and ECG traits in both directions. The outcomes will help us understand why individuals with schizophrenia are at increased risk of sudden cardiac death – knowledge which is crucial to improve life expectancy in this vulnerable population.

## Methods

We leveraged summary-level data from the largest available GWASs and applied various genetics-based methods displayed in **Figure 1**. The primary measure of interest, schizophrenia diagnosis, was chosen because it is the psychiatric disorder linked most consistently and strongly to cardiovascular disease and mortality. Schizophrenia cases had a clinical diagnosis within the schizophrenia spectrum disorder, based on the widely accepted DSM-IV criteria.^34^ Information on the measurement of the two arrhythmic disorders (atrial fibrillation^32^ and Brugada syndrome^29^) and ECG traits (heart rate during activity^35^, heart rate recovery after activity^35^, heart rate variability^33^, QT interval^30^, PR^31^, JT^30^, and QRS^30^) can be found in **Table 1**.

**Table 1.** Overview of genome-wide association studies (GWASs) that were used to conduct genetics-based analytical methods.

| Phenotype | GWAS reference | Measurement | Sample size | $h^2_{SNP}$ |
| --- | --- | --- | --- | --- |
| Schizophrenia | Trubetskoy et al., 2022 | Cases were individuals diagnosed with a schizophrenia spectrum disorder, based on DSM-IV criteria. | 53,386 cases<br>77,258 controls | 0.21 (se 0.007) |
| Atrial fibrillation (AF) | Roselli et al., 2018 | Cases were individuals diagnosed with paroxysmal or permanent AF, or with atrial flutter. | 55,114 cases<br>482,295 controls | 0.14 (se 0.013) |
| Brugada syndrome | Barc et al., 2022 | Cases were individuals diagnosed with a type 1 Brugada ECG, defined as a coved type ST elevation at baseline (spontaneous) or after a drug challenge test, in one or more leads in the right precordial leads V1 and/or V2 in the standard position (4th intercostal space) or in high positions (2nd or 3rd intercostal spaces). The diagnoses were made by a cardiac electrophysiologist with expertise in Brugada syndrome. | 2,820 cases<br>10,001 controls | 0.19 (se 0.037) |
| HR activity | Ramírez et al., 2018 | Automated heart rate (HR) measurements and electrocardiogram (ECG) recordings from individuals who participated in the ‘Cardio test’ of the UK Biobank (UKB) study were analysed. HR activity reflects HR measured in beats per minute during exercise (cycling). | 66,811 | 0.15 (se 0.011) |
| HR recovery | Ramírez et al., 2018 | Automated heart rate (HR) measurements and electrocardiogram (ECG) recordings from individuals who participated in the ‘Cardio test’ of the UK Biobank (UKB) study were analysed. HR activity reflects HR measured in beats per minute 1 minute post-exercise (cycling). | 66,678 | 0.12 (se 0.014) |
| HR variability | Nolte et al., 2017 | The root mean square of the successive differences of inter beat intervals (RMSSD), was computed, which reflects HR variability. | 26,523 to 46,952* | 0.11 (se 0.028) |
| PR interval | Ntalla et al., 2020 | The PR interval (in milliseconds) was determined based on ECG recordings and reflects conduction from the atria to ventricles, across specialized conduction tissues such as the atrioventricular node and the His-Purkinje system. Individuals were excluded in case of: extreme PR interval values (320 ms), second/third degree heart block, AF on the ECG, or a history of myocardial infarction or heart failure, Wolff–Parkinson–White syndrome, a pacemaker, receiving class I and class III antiarrhythmic medications, digoxin, and pregnancy. | 293,051 | 0.62** (se 0.064) |
| QT interval | Young et al., 2022 | The QT interval (in milliseconds) was determined based on ECG recordings and represents the sum of ventricular depolarization (QRS duration) and repolarization (JT interval). Individuals were excluded in case of: prevalent myocardial infarction or heart failure, pregnancy at the time of recruitment, implantation of a pacemaker or implantable cardiac defibrillator, QRS duration >120 ms, or right or left bundle branch block or atrial fibrillation on ECG. | 252,977 | 0.17 (se 0.034) |
| JT interval | Young et al., 2022 | The JT interval (in milliseconds) was determined based on ECG recordings and represents repolarization at an organ level. Individuals were excluded in case of: prevalent myocardial infarction or heart failure, pregnancy at the time of recruitment, implantation of a pacemaker or implantable cardiac defibrillator, QRS duration >120 ms, or right or left bundle branch block or atrial fibrillation on ECG. | 252,977 | 0.16 (se 0.031) |
| QRS duration | Young et al., 2022 | QRS duration (in milliseconds) was determined based on ECG recordings and represents ventricular depolarization. Individuals were excluded in case of: prevalent myocardial infarction or heart failure, pregnancy at the time of recruitment, implantation of a pacemaker or implantable cardiac defibrillator, QRS duration >120 ms, or right or left bundle branch block or AF on ECG | 252,977 | 0.12 (se 0.014) |

\*Only SNPs that were genome-wide significant ( $p < 5E-08$ ) were analyzed in the larger sample of 46,952 individuals. $h^2_{SNP}$ =
\*\*Note that this SNP-based heritability estimate is considerably higher than what is reported in the original GWAS of Ntalla et al (2020) due to a difference in the selection criteria for which SNPs to include for Linkage Disequilibrium Score Regression analysis within our own pipeline

**Figure 1.**
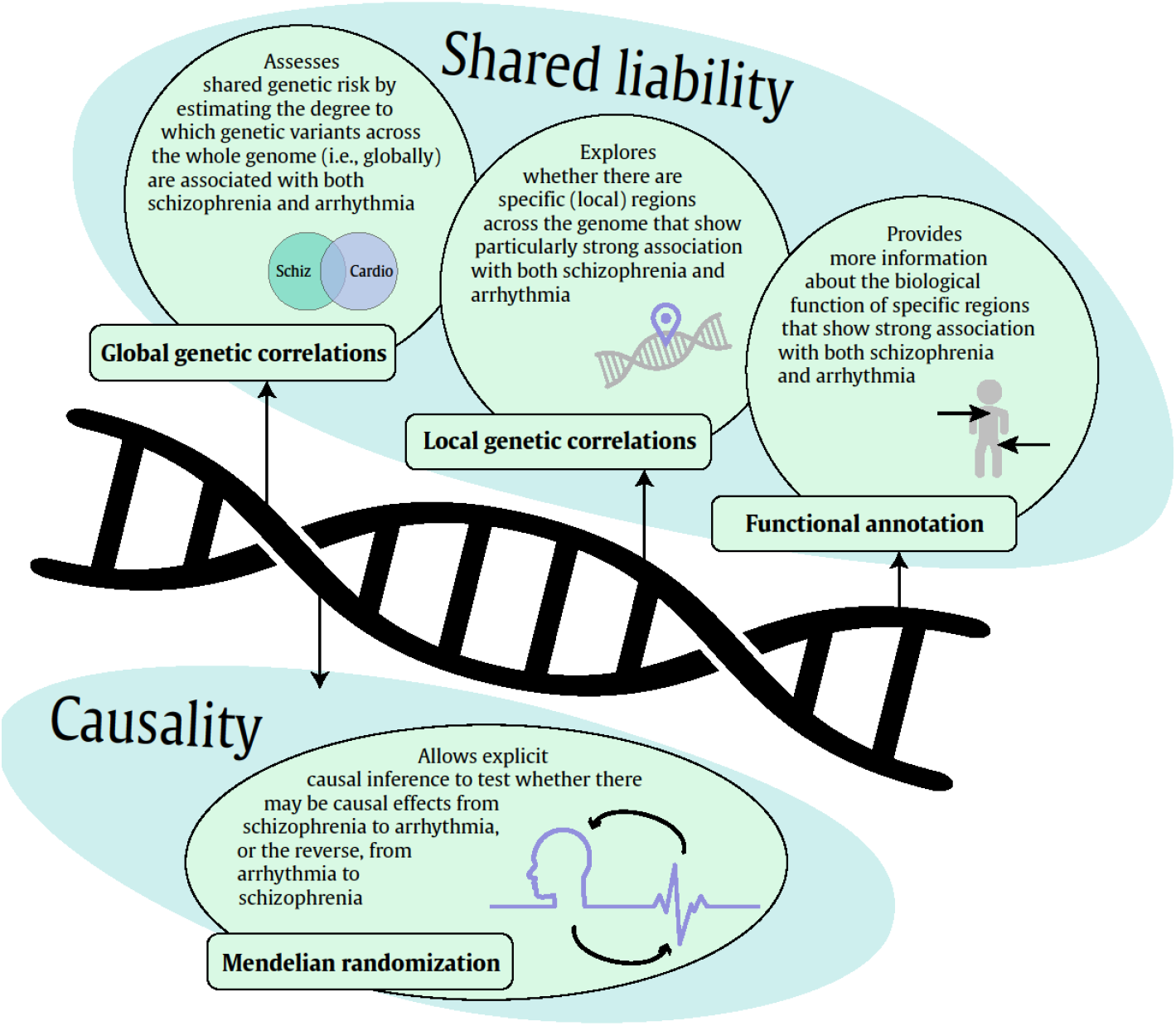
Overview of the genetics-based methods that were applied to investigate the mechanisms of schizophrenia with arrhythmic disorders and ECG traits. First, we examined whether there are shared genetic risk factors that overlap between schizophrenia and arrhythmic disorders and ECG traits, by estimating global and local genetic correlations. For regions of the genome that show a correlation between schizophrenia and cardiac functioning, we ran a range of functional annotation analyses to better understand the biological mechanisms involved. Subsequently, we applied bidirectional MR to investigate causal associations between schizophrenia and cardiac function.

### Global genetic correlations

To estimate genome-wide genetic correlations we applied Linkage Disequilibrium (LD) Score regression using SNP effect estimates from the existing GWASs.^21^ We first filtered the GWAS summary statistics by excluding SNPs with a minor allele frequency (MAF)<0·01, missing values and infinite test statistic values. Next, we extracted SNPs available in the HapMap 3 reference panel. For each trait-pair, genetic covariance was estimated using the slope from the regression of the product of z-scores from the two corresponding GWASs on the LD score. A global genetic correlation represents the genetic covariation between two traits based on all polygenic effects captured by the SNPs included in the GWASs. LD scores were based on the HapMap 3 reference panel (European). In order to establish whether the strength of genetic correlation varies by SNP variant frequency, for which there is some evidence^23^, we also computed MAF-stratified genetic correlations. We created strata of MAF between boundary values 0·05, 0·11, 0·22, 0·35, and 0·50, consistent with Perry et al., 2022.^23^

### Local genetic correlations

We used LAVA (Local Analysis of [co]Variant Association) to assess local genetic correlations of schizophrenia with arrhythmic disorders and ECG traits.^24^ A total of 2495 pre-defined regions across the entire genome were assessed. These regions were provided alongside the software and were created by partitioning the genome into blocks of approximately equal size (∼1 Mb) while minimizing LD between them. The 1000 Genomes European panel (MAF>0·01) was used as a reference panel. Only regions that showed a highly significant univariate heritability (*h*^2^_*SNP*_ *p*<0·0001) for both schizophrenia and the cardiac trait were tested for local genetic correlation. Per schizophrenia-cardiac trait combination, an FDR (False Discovery Rate) correction for multiple testing was applied to adjust for the number of tested regions.

### Functional annotation

For regions that showed (FDR-corrected) evidence of a schizophrenia-cardiac trait correlation, we performed functional annotation using FUMA (Functional Mapping and Annotation of GWASs).^25^ We separately investigated regions with a positive or negative genetic correlation, because enrichment in positively versus negatively associated regions would have a different interpretation. For instance, enrichment in a negative region could suggest opposing underlying biological pathways. We created lists of ‘positive’ and ‘negative’ genes for each trait pair by looking up all protein coding genes that fell within the associated regions according to the NCBI (National Center for Biotechnology Information) reference data.^36^ These lists were then annotated using the FUMA GENE2FUNC module, excluding the HLA region.^25^ First, we assessed with which traits these genes had previously been found to associate in the GWAS catalogue. Second, we assessed biological processes underlying the associations through Gene Ontology (GO:0050896) gene set enrichment analysis, i.e. by assessing whether the genes were overrepresented in predefined gene sets. Finally, we assessed evidence for expression of these genes in the 30 available tissue types of the GTEx (Genotype-Tissue Expression) project (v8^37^). Specifically, we assessed if genes in regions with a significant schizophrenia-cardiac trait association were differentially (more or less) expressed in a tissue, as compared to all the other tissues. Differential tissue expression can provide clues to the location of the biological processes driving the genetic association between schizophrenia and cardiac traits.^38^ In the main results, we focus on enrichment in positively associated regions, because these showed stronger and more uniform enrichment patterns and have a more straightforward interpretation.

### Causal inference with Mendelian randomization

We applied MR to assess evidence for causal effects of liability to schizophrenia on arrhythmic disorders and ECG traits as well as of arrhythmic disorders and ECG traits on schizophrenia risk. For an MR analysis to be valid, the genetic variants selected as instruments should 1) associate robustly and strongly with the exposure, 2) be independent of confounders, and 3) not directly influence the outcome, except through their effect on the exposure.^39^ If these assumptions are met, the causal effect of the exposure on the outcome can be estimated with inverse-variance weighted regression (IVW).^40^ While IVW provides an indication of causality, it presumes that all assumptions are met, which is unlikely for complex traits. The most important source of potential bias is horizontal pleiotropy; SNPs affecting the outcome without going through the exposure. To verify results obtained from IVW, we applied five sensitivity methods. If a finding is consistent across these methods, it constitutes robust evidence for causality. Due to inherently lower power of the sensitivity methods, some decrease in the strength of statistical evidence (but not the effect size) is expected even for a true causal effect.

The sensitivity methods we applied are: Weighted Median regression, which provides a consistent estimate of a causal effect, even when <50% of the weight of the instrument does not satisfy the MR assumptions^41^; Weighted Mode regression, which can provide a consistent estimate of a causal effect if the most frequent SNP-effects are contributed by valid SNPs^42^; MR-Egger, which can explicitly test for horizontal pleiotropy by freely estimating an intercept (instead of fixing it at zero) that captures the average horizontally pleiotropic effect^43^; MR-PRESSO (MR pleiotropy residual sum and outlier), which assesses horizontal pleiotropy (global test), corrects for it by removing outliers and evaluates differences in the estimate of the causal effect before and after removal of outliers (distortion test)^44^; Steiger filtering, which explicitly corrects for reverse causality by identifying and then excluding SNPs that explain a larger amount of variance in the outcome, compared to the exposure.^45^ We also computed Cochran’s Q to assess heterogeneity between SNP-estimates in each instrument and for potentially causal findings we performed leave-one-out IVW and displayed all SNP-estimates in a funnel plot to assess (a)symmetry. To assess instrument strength, we computed the F-statistic (F>10 is sufficiently strong). All MR analyses were conducted in R (4.2.0), using packages: “*TwoSampleMR,” “GSMR,” “psych”* and “*MR-PRESSO*”.

## Results

### Global genetic correlations

Global genetic correlations, based on all SNPs included in the GWASs, as well as MAF-stratified genetic correlations are presented in **Table 2**. Evidence for (modest) global genetic correlation (r_g_=0·14, 95% CIs=0·06 to 0·21, *p=*4·0E-04) was only present for schizophrenia and Brugada syndrome. When stratifying on MAF, there was some indication of stronger correlation for the lower compared to the higher MAF strata, but differences were minor.

Using LAVA, we found a picture of local correlations across the genome, both in the positive and negative direction (**Figure 2, Table S1**). After filtering on univariate heritability, between 105 and 264 regions per schizophrenia-cardiac trait combination were tested for local genetic correlation, resulting in between 20 and 60 nominally significantly associated regions per trait combination. Of particular interest are the local correlations that survived FDR-correction. For all trait combinations there were 4 (SCZ-AF & SCZ-HRV) to 33 (SCZ-QT) regions with significant signal after correction. For most trait pairs, there were both regions with positive and regions with negative correlation. To assess how these local correlations relate to the genome-wide significant loci of the original GWASs, we created Miami plots of the original GWAS SNP-estimates for each schizophrenia-cardiac trait pair and identified the SNPs in the local regions that showed significant correlation (**Figures S1-S10)**.

**Figure 2.**
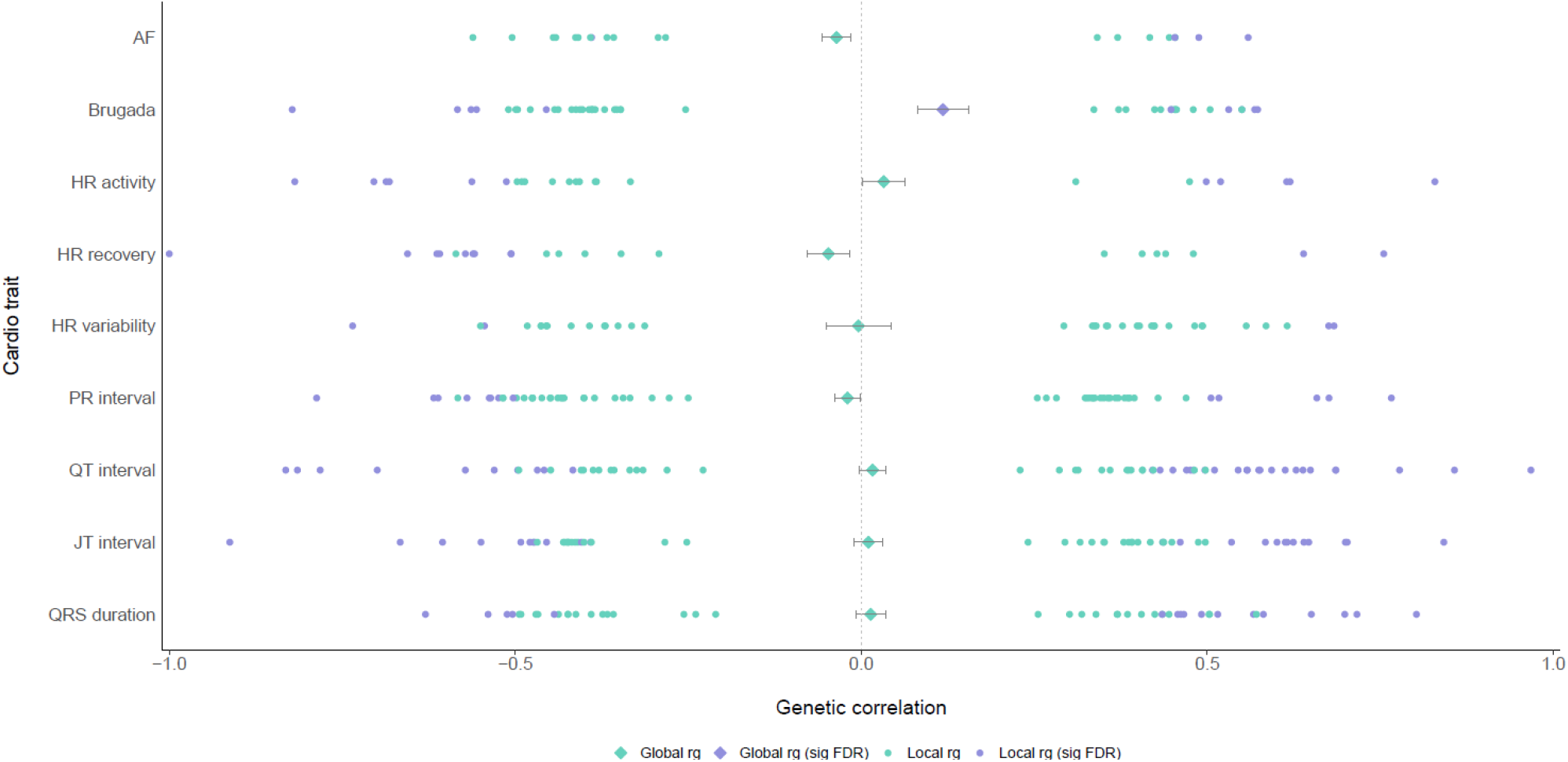
Results of global and local genetic correlation analyses between schizophrenia and two arrhythmic disorders and seven ECG traits. The global genetic correlations, computed with Linkage Disequilibrium score regression analyses including all Single Nucleotide Polymorphisms in the respective genome-wide association studies, are shown as diamonds in the middle. Local significant genetic correlations for genomic regions computed with LAVA (Local Analysis of [co]Variant Association) are shown as dots, with each dot representing a region comprising a couple of thousands Single Nucleotide Polymorphisms.

### Functional annotation of shared genomic regions

To obtain a better understanding of the biological significance of the shared genomic regions, we performed functional annotation analysis, looking separately at genes in regions with positive or negative schizophrenia-cardiac trait associations.

The identified gene-sets were found to be associated with many traits in the GWAS catalogue. Genes in regions with positive schizophrenia-QT and schizophrenia-JT associations were associated with auto-inflammatory and immune-related traits (**Fig S11A**). Genes in regions with negative schizophrenia-JT and schizophrenia-PR associations were mostly associated with metabolic traits (**Fig S11B**).

Enrichment in GO gene sets for biological processes was found mainly for genes in regions with a positive association between schizophrenia and Brugada syndrome (**Figure 3A & Fig S12**). These were genes were mostly related to viral response mechanisms and immune-related processes. We did not observe enrichment in the case of other trait combinations, with the exception of four terms for genes in regions that showed a negative association of schizophrenia with HR reactivity and QT duration (**Figure S12**). Including the HLA region yielded consistent results and additional enrichment in immune-related GO terms for genes in positive schizophrenia-Brugada and schizophrenia-QT regions (results not shown).

**Figure 3.**
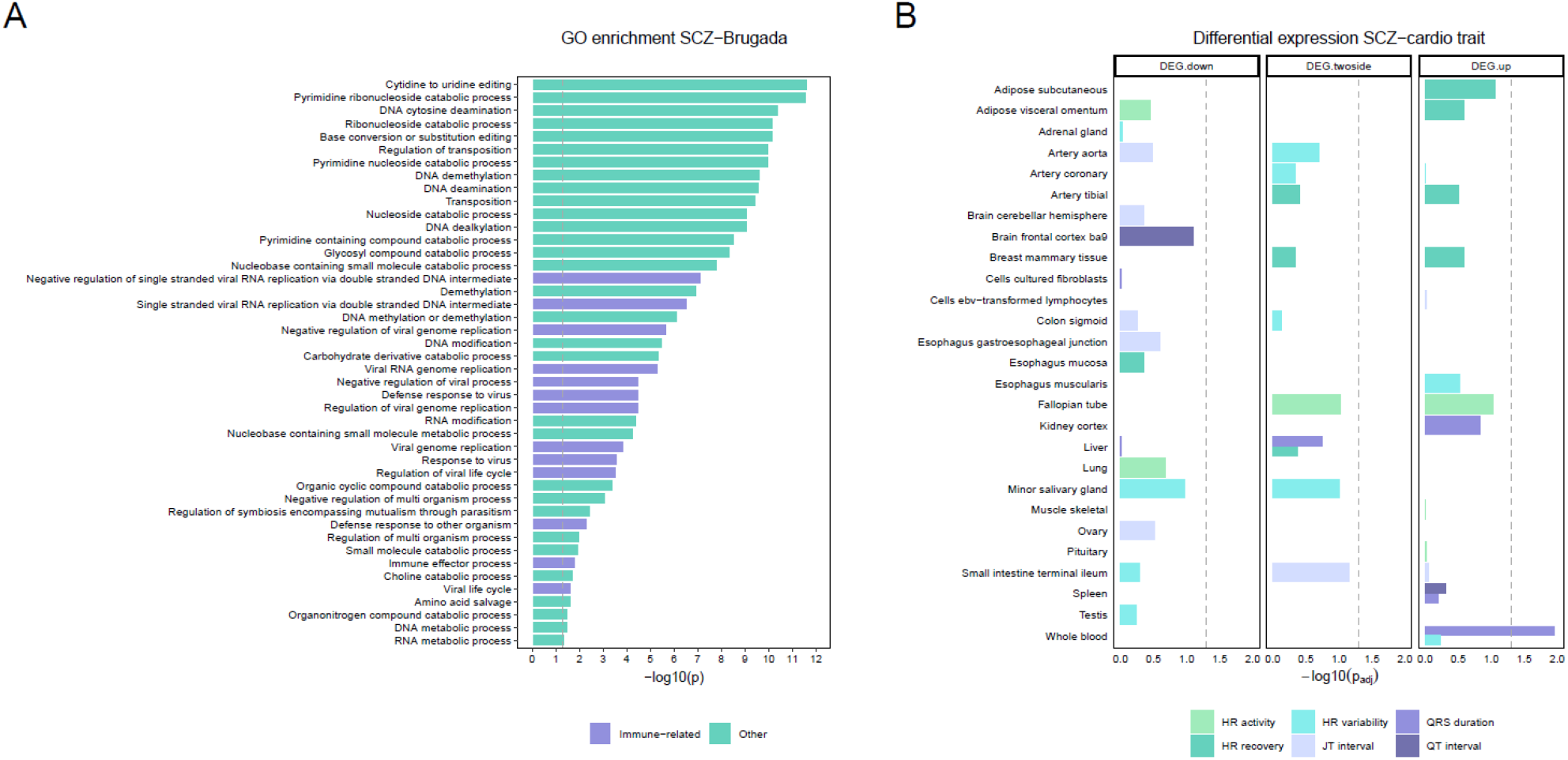
Functional significance of genes in regions that correlate positively between schizophrenia and an arrhythmic disorder or ECG trait. **Panel A** shows significant enrichment (after FDR-correction) in biological process GO terms for all genes in regions with a positive genetic correlation between schizophrenia and Brugada. GO terms that are primarily related to viral response and immune processes are highlighted in purple. **Panel B** shows differential tissue expression (downregulation, upregulation, and combined, at marginal *p<*.*05*) for genes in regions that showed a positive genetic correlation between a schizophrenia and a cardiac function trait pair. The x-axes in both panels show the log-transformed p-value after FDR-correction, with the dashed lines indicating the significance threshold (*p*_*FDR*_*<*.*05*).

**Figure 3B** shows differential expression across the 30 available tissue types of the GTEx (Genotype-Tissue Expression) project for positive trait pair regions (marginal *p<*.05). After FDR-correction, there was only one significant finding for the positive trait pair regions; genes in regions with positive associations between schizophrenia and QRS duration were upregulated (expressed at higher levels) in whole blood. This means that genes shared between schizophrenia and QRS duration are more expressed in whole blood as compared to other tissues, suggesting that a biological process within this tissue drives the association. Artery (aorta, coronary, tibial) and two brain regions were among the tissues showing marginal enrichment. For the negative trait pairs there was less differential expression and none of the tissues survived correction for multiple testing (full results in **Figure S13**).

### Causal effects between schizophrenia and arrhythmic disorders and ECG traits

Results of bidirectional MR analyses between liability to schizophrenia and cardiac traits are shown in **Figure 4**. There was strong evidence for a causal, increasing effect of liability to schizophrenia on Brugada syndrome risk (IVW OR=1·15, 95% CIs=1·03 to 1·28, *p=*0·009), which was consistent in effect size across a range of sensitivity methods (for scatterplot, funnel plot, and leave-one-out analyses see **Figures S14-S19**). The direction of causality was confirmed by Steiger. MR-Egger provided good evidence for causality (OR=1·67, 95% CIs=1·08 to 2·56, *p=*0·022). While there was strong evidence for heterogeneity between the different SNP-effects (Cochran’s Q, *p*=1.2E-04; **Table S2**), there was no indication for horizontal pleiotropy (Egger intercept=-0·03, *p=*0·104). There was also evidence for a causal, increasing effect of liability to schizophrenia on heart rate during activity (IVW beta=0·25, 95% CIs=0·05 to 0·45, *p=*0·015) consistent across sensitivity methods. Although there was weak evidence for horizontal pleiotropy (MR-Egger intercept=-0·05, 95% CIs=-0·10 to 0·00, *p=*0·073), the MR-Egger slope still showed evidence for causality (beta=0·97, 95% CIs=0·23 to 1·71, *p=*0·011). There was no evidence for causality for any other relationship.

**Figure 4.**
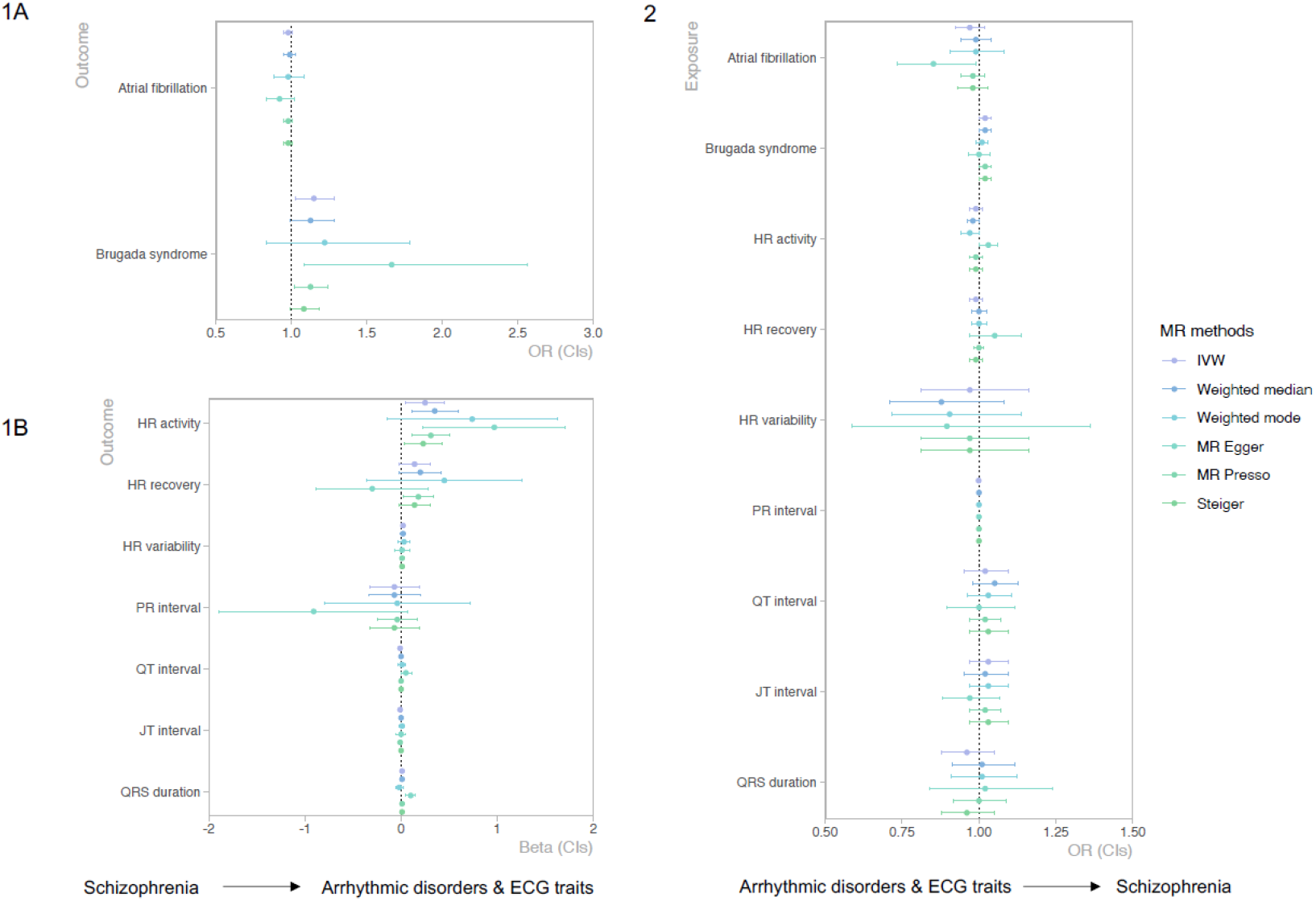
Bidirectional Mendelian randomization analyses from liability to schizophrenia onto arrhythmic disorders and ECG traits and vice versa, from arrhythmic disorders and ECG traits to schizophrenia risk. Note that the Inverse Variance Weighted (IVW) analysis is the main analytical method and all other analyses should be seen as sensitivity methods to check whether any potential causal effect indicated by IVW holds (i.e., if there is a significant result for one of the sensitivity methods but not for the IVW, we would not consider that evidence for causality). MR-egger slope indicates the estimated causal effect, while the MR-Egger intercept reflects horizontal pleiotropy (if the p-value for the intercept is significant this indicates that there is horizontal pleiotropy present). The I-squared statistic, which assesses whether the NOME assumption was satisfied and an MR-Egger analysis can be considered reliable, ranged between acceptable to very good values (0.60 and 0·98), if I-squared was <0·90, Egger SIMEX (simulation extrapolation) was applied to correct for any potential bias.

To better understand the pathway through which schizophrenia may causally increase Brugada syndrome risk, we employed multivariable MR to add each of the heart rate and ECG traits. The main effect of liability to schizophrenia on Brugada stayed consistent (**Table S3**), suggesting that these cardiac parameters do not drive the causal relationship.

## Discussion

This study is the first to comprehensively investigate the relation of schizophrenia with arrhythmic disorders and ECG traits using advanced genetics-based methods. We found evidence for modest global genetic correlation between schizophrenia and Brugada syndrome, but no evidence for global genetic correlations between schizophrenia and eight other traits (atrial fibrillation, heart rate during activity and recovery, heart rate variability, PR interval, QT interval, JT interval, and QRS duration). When considering specific regions across the genome, a pattern of widespread local genetic correlations, both negative and positive, emerged for all trait pairs. Functional annotation showed that the genes located in regions that correlated between schizophrenia and Brugada syndrome were mainly involved in immune-related processes and viral response mechanisms. Finally, Mendelian randomization showed strong evidence for causal, increasing effects of liability to schizophrenia on Brugada syndrome and heart rate during physical activity.

The lack of evidence for (strong) global genetic correlation concurs with previous studies that found similarly low genetic correlations between schizophrenia and different cardio-vascular and cardio-metabolic traits.^22,23^ We did find significant correlations between schizophrenia and all cardiac traits (both positive and negative) for specific genomic regions, indicating that a global correlation overlooks important local processes by averaging out opposing effects. Functional annotation showed that the regions that correlated significantly were largely enriched in genes related to the immune-system, suggesting that schizophrenia and arrhythmia share common immunological pathways. These findings are in line with an increasing body of literature suggesting a shared immunological aetiology between cardio-metabolic traits and serious mental illness, such as major depressive disorder.^46^ The strongest evidence was found for regions correlating positively between schizophrenia and Brugada syndrome, which were particularly enriched for viral-response pathways. This concurs with the theory that a viral infection in mothers during pregnancy increases the risk of schizophrenia in offspring.^47,48^ The role of the immune system in the aetiology of Brugada is unclear and should be studied further.^49^

Another striking finding was the causal, increasing effect of liability to schizophrenia on Brugada syndrome. The pathophysiology of Brugada syndrome involves dysfunction of ion, primarily sodium, channels.^15^ Interestingly, we previously showed evidence for a causal effect of liability to schizophrenia on early repolarization, an ECG pattern which is, like Brugada, linked to increased risk of sudden cardiac death and suspected to involve ion channel dysfunction.^22^ These findings indicate that schizophrenia increases the risk of such arrhythmias, but the exact biological pathway remains unclear. It could be that when there is already a high liability for Brugada syndrome, an ongoing psychotic state acts as a catalyst.^50^ Some people start off with normal ECG readings after which factors such as fever or metabolic disorders ‘unmask’ a Brugada pattern and schizophrenia may be another such factor.^14^ Dysfunction of the autonomic nervous system might play a role herein, as it is involved in schizophrenia and possibly also Brugada syndrome.^26^ It is not likely that the effects we found are due to psychotropic medication use, since such medication primarily impacts the QT interval and the effect of schizophrenia on Brugada was not mediated by QT in a multivariable MR analysis. Importantly, our findings suggest that systematic screening for Brugada syndrome among patients with schizophrenia is warranted. Since some people with Brugada syndrome are asymptomatic and the preventive treatment of placing an ICD (Implantable Cardioverter-Defibrillator) is invasive,^51^ further research should focus on identifying patients with Brugada who are at increased risk of sudden cardiac death. This is particularly important for those with schizophrenia, as they are already at increased risk for cardiovascular disease and mortality, even without Brugada syndrome.^52^

The current study uniquely used advanced methods and large, powerful genetic samples to study relations between (rare) complex disorders. The novel biological pathways that we report can lead to important unexplored avenues of research. Besides these important strengths, there are also limitations to consider. The serious nature of schizophrenia means that those who suffer most may not have been able to participate in research, causing selection bias.^53,54^ For MR in particular, assortative mating, dynastic effects and residual population stratification may have caused bias, for which we were not able to correct without the availability of large family samples.^55^ Another limitation is that well-powered datasets for ancestries other than European were not available, limiting generalizability. Such bias is widespread in medical and genetics research.^56^

In sum, we report limited global genetic overlap, but widespread local genetic correlations of schizophrenia with arrhythmic disorders and ECG traits. We highlighted specific biological mechanisms that may be responsible for local shared aetiology, with immunological and viral response processes emerging as important candidates for follow-up research. There was highly robust evidence for a causal effect of liability to schizophrenia on Brugada syndrome, building on recent genetically informed studies that indicated effects of schizophrenia on heart failure as well as functional measures such as decreased cardiac volumes.^22,57^ Overall, our findings emphasize that cardiac monitoring needs to be performed more frequently among individuals with schizophrenia than is currently done, and that treatment of both psychosis and cardiac abnormalities should be started timely in order to decrease mortality in this vulnerable population.^58^

## Data Availability

The findings of this study are based on summary-level data of published GWAS, all of which are publicly available or can be requested from the corresponding author, JLT.

## Acknowledgements

JLT is supported by a European Research Council (ERC) Starting grant (UNRAVEL-CAUSALITY, grant number 101076686). KJHV and JLT are supported by Foundation Volksbond Rotterdam. CRB is supported by the Dutch Heart Foundation (PREDICT2 project, CVON 2018-30) and the Leducq Foundation (project 17CVD02). JAP was supported by the US National Institutes of Mental Health (R01MH123724). JAP and JBerg were supported by European Union’s Horizon 2020 Research and Innovation Programme (CoMorMent project; Grant #847776). RT is supported by the Canada Research Chairs program.

## Authors contributions

JLT, CRB, RT, and KJHV conceived of the initial research idea and analysis plan. JLT conducted the Mendelian randomization analyses, ABT and DJM conducted the genetic correlation analyses, JBerg and JP conducted the functional follow up analyses. JLT wrote and finalized the manuscript, with important contributions from all the other authors (ABT, DJM, RT, RRV, DD, JMV, JBerg, JBarc, JAP, CRB, KJHV). ABT, RRV, JBerg and JAP created the main figures in the paper, crucial to clarify the aims and results of the study. All of the authors read and reviewed the final version of the manuscript.

## Data sharing

No new data were collected or created for the current study, all analysis were based on summary-level data of existing GWASs and biobank studies. The analysis plans of the current study were pre-registered at OSF (https://osf.io/fe4ms).

