## Supplementary material for "Associations of schizophrenia with arrhythmic disorders and electrocardiogram traits: an in-depth genetic exploration of population samples"

1. **Complete reference list main publication**	2-4
2. **Local genetic correlation analyses**
a. Supplementary Figure S1 (overlap in locally correlating regions across trait-pairs)	5
b. Supplementary Figures S2 to 10 (Manhattan plots with locally correlating regions)	6-10
c. Supplementary Table S1 (global and MAF-stratified genetic correlations)	11
d. Supplementary Table S2 (list of locally correlating regions)	12-14
3. **Functional annotation analyses**
a. Supplementary Figure S11 (GWAS catalogue phenotypes)	15
b. Supplementary Figure S12 (GO biological processes enrichment)	16
c. Supplementary Figure S13 (Differential tissue expression)	17
4. **Mendelian randomization analyses**
a. Supplementary Figures S14 to 15 (scatter plots causal findings)	18
b. Supplementary Figures S16 to 17 (funnel plots causal findings)	19
c. Supplementary Figures S18 to 19 (leave-one-out analyses causal findings)	20-21
d. Supplementary Table S3 (Cochran’s Q statistic to assess heterogeneity)	22
e. Supplementary Table S4 (Multivariable MR analyses)	23
1. **Complete reference list main publication**

31	Ntalla I, Weng L-C, Cartwright JH, *et al.* Multi-ancestry GWAS of the electrocardiographic PR interval identifies 202 loci underlying cardiac conduction. *Nat Commun* 2020; **11**: 2542.

32	Roselli C, Chaffin MD, Weng L-C, *et al.* Multi-ethnic genome-wide association study for atrial fibrillation. *Nat Genet* 2018; **50**: 1225–33.

33	Nolte IM, Munoz ML, Tragante V, *et al.* Genetic loci associated with heart rate variability and their effects on cardiac disease risk. *Nat Commun* 2017; **8**: 15805.

34	Trubetskoy V, Pardiñas AF, Qi T, *et al.* Mapping genomic loci implicates genes and synaptic biology in schizophrenia. *Nature* 2022; : 1–13.

35	Ramírez J, Duijvenboden S van, Ntalla I, *et al.* Thirty loci identified for heart rate response to exercise and recovery implicate autonomic nervous system. *Nat Commun* 2018; **9**: 1947.

36	Maglott D, Ostell J, Pruitt KD, Tatusova T. Entrez Gene: gene-centered information at NCBI. *Nucleic Acids Research* 2005; **33**: D54–8.

37	THE GTEX CONSORTIUM. The GTEx Consortium atlas of genetic regulatory effects across human tissues. *Science* 2020; **369**: 1318–30.

38	Ashburner M, Ball CA, Blake JA, *et al.* Gene Ontology: tool for the unification of biology. *Nat Genet* 2000; **25**: 25–9.

39	Skrivankova VW, Richmond RC, Woolf BAR, *et al.* Strengthening the reporting of observational studies in epidemiology using mendelian randomisation (STROBE-MR): explanation and elaboration. *BMJ* 2021; **375**: n2233.

40	Davies NM, Holmes MV, Davey Smith G. Reading Mendelian randomisation studies: a guide, glossary, and checklist for clinicians. *BMJ* 2018; **362**: k601.

41	Bowden J, Davey Smith G, Haycock PC, Burgess S. Consistent Estimation in Mendelian Randomization with Some Invalid Instruments Using a Weighted Median Estimator. *Genet Epidemiol* 2016; **40**: 304–14.

42	Hartwig FP, Davey Smith G, Bowden J. Robust inference in summary data Mendelian randomization via the zero modal pleiotropy assumption. *Int J Epidemiol* 2017; **46**: 1985–98.

43	Bowden J, Davey Smith G, Burgess S. Mendelian randomization with invalid instruments: effect estimation and bias detection through Egger regression. *Int J Epidemiol* 2015; **44**: 512–25.

44	Verbanck M, Chen C-Y, Neale B, Do R. Detection of widespread horizontal pleiotropy in causal relationships inferred from Mendelian randomization between complex traits and diseases. *Nat Genet* 2018; **50**: 693–8.

45	Hemani G, Tilling K, Smith GD. Orienting the causal relationship between imprecisely measured traits using GWAS summary data. *PLOS Genetics* 2017; **13**: e1007081.

46	Milaneschi Y, Lamers F, Berk M, Penninx BWJH. Depression Heterogeneity and Its Biological Underpinnings: Toward Immunometabolic Depression. *Biol Psychiatry* 2020; **88**: 369–80.

47	Børglum AD, Demontis D, Grove J, *et al.* Genome-wide study of association and interaction with maternal cytomegalovirus infection suggests new schizophrenia loci. *Mol Psychiatry* 2014; **19**: 325–33.

48	Robinson N, Ploner A, Leone M, Lichtenstein P, Kendler KS, Bergen SE. Impact of Early-Life Factors on Risk for Schizophrenia and Bipolar Disorder. *Schizophrenia Bulletin* 2023; **49**: 768–77.

49	Oliva A, Grassi S, Pinchi V, *et al.* Structural Heart Alterations in Brugada Syndrome: Is it Really a Channelopathy? A Systematic Review. *Journal of Clinical Medicine* 2022; **11**: 4406.

50	Bär K-J. Cardiac Autonomic Dysfunction in Patients with Schizophrenia and Their Healthy Relatives – A Small Review. *Frontiers in Neurology* 2015; **6**. https://www.frontiersin.org/articles/10.3389/fneur.2015.00139 (accessed April 3, 2023).

51	Popa IP, Șerban DN, Mărănducă MA, Șerban IL, Tamba BI, Tudorancea I. Brugada Syndrome: From Molecular Mechanisms and Genetics to Risk Stratification. *Int J Mol Sci* 2023; **24**: 3328.

52	Goldfarb M, De HM, Detraux J, *et al.* Severe Mental Illness and Cardiovascular Disease. *Journal of the American College of Cardiology* 2022; **80**: 918–33.

53	Martin J, Tilling K, Hubbard L, *et al.* Association of Genetic Risk for Schizophrenia With Nonparticipation Over Time in a Population-Based Cohort Study. *Am J Epidemiol* 2016; **183**: 1149–58.

54	Tyrrell J, Zheng J, Beaumont R, *et al.* Genetic predictors of participation in optional components of UK Biobank. *Nat Commun* 2021; **12**: 886.

55	Brumpton B, Sanderson E, Heilbron K, *et al.* Avoiding dynastic, assortative mating, and population stratification biases in Mendelian randomization through within-family analyses. *Nat Commun* 2020; **11**: 3519.

56	Abdellaoui A, Verweij KJH. Dissecting polygenic signals from genome-wide association studies on human behaviour. *Nat Hum Behav* 2021; **5**: 686–94.

57	Pillinger T, Osimo EF, Marvao A de, *et al.* Effect of polygenic risk for schizophrenia on cardiac structure and function: a UK Biobank observational study. *The Lancet Psychiatry* 2023; **10**: 98–107.

58	Nielsen RE, Banner J, Jensen SE. Cardiovascular disease in patients with severe mental illness. *Nat Rev Cardiol* 2021; **18**: 136–45.

2. **Local genetic correlation analyses**

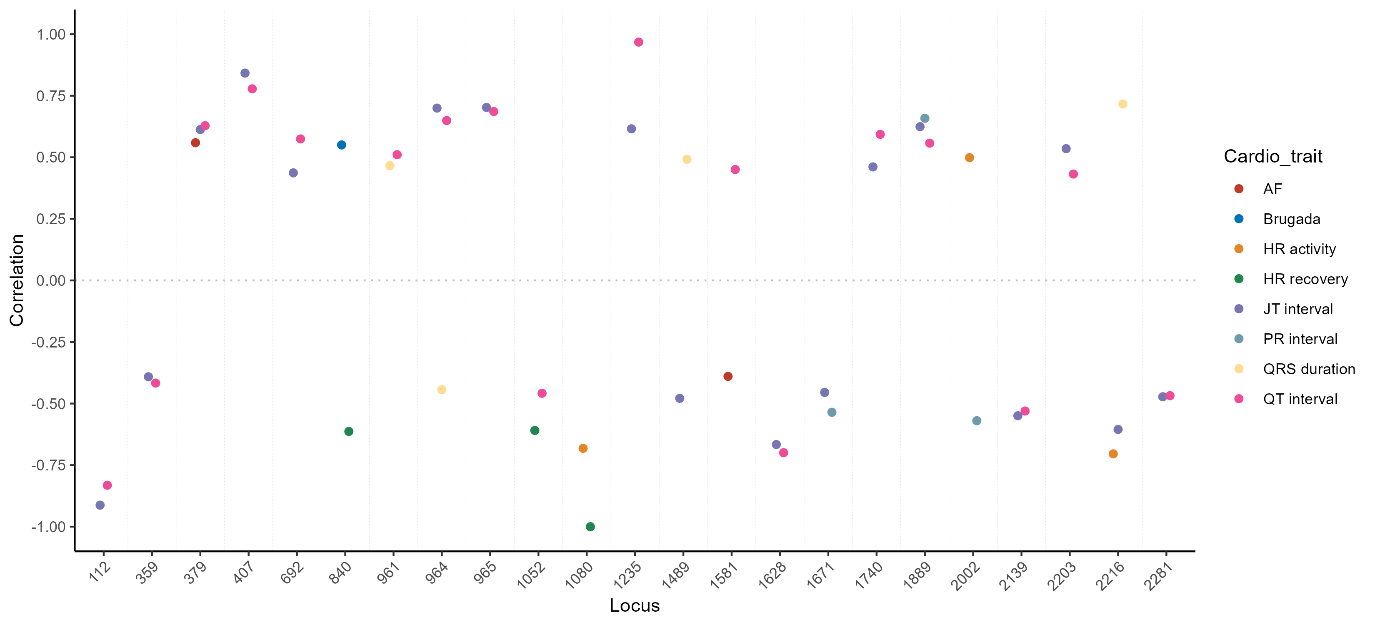

**Figure S1***.* Regions that emerged in two or more cardio trait – schizophrenia local correlation analyses are plotted. The x-axis shows the 23 regions that overlapped between LAVA analyses and the y-axis shows the local genetic correlation between the cardio traits and schizophrenia for the involved regions.

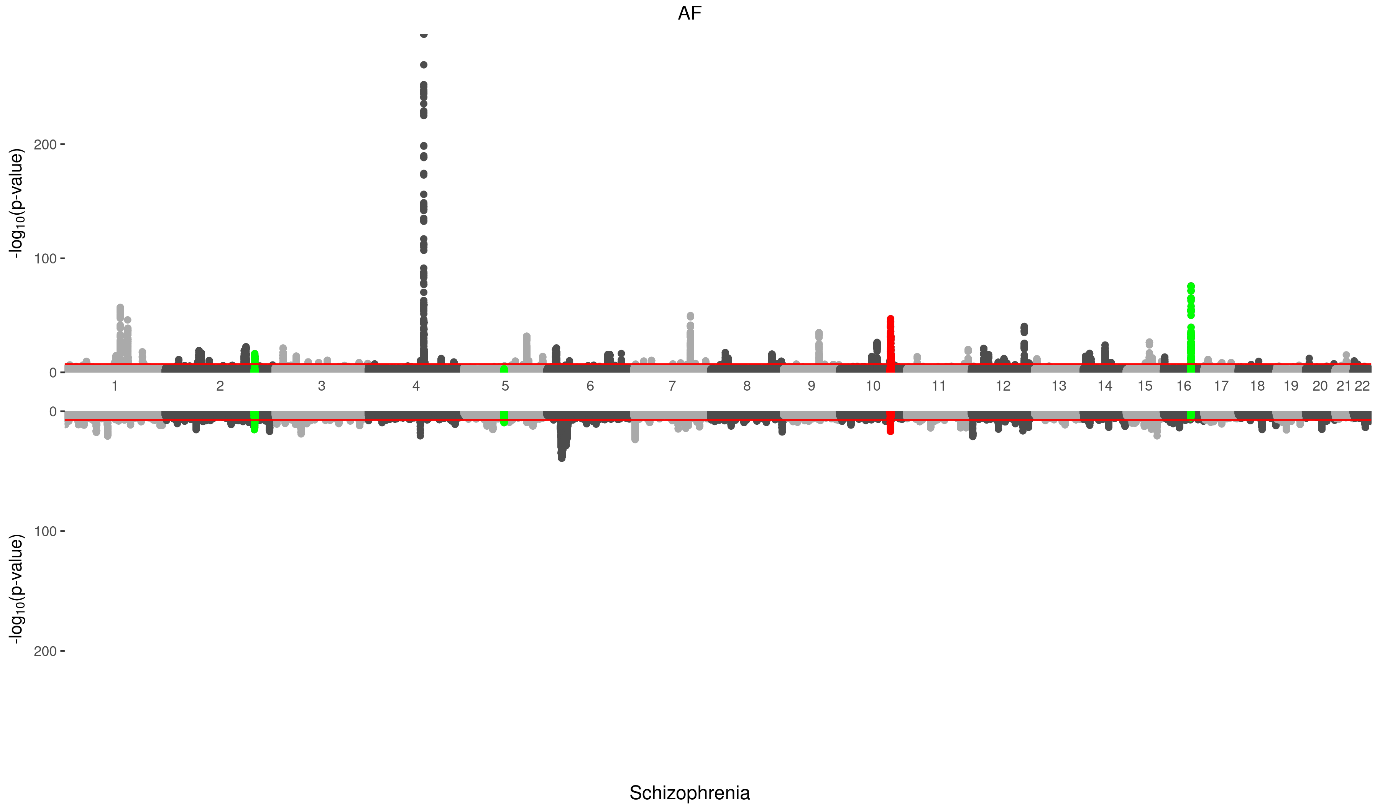

**Figure S2.** Miami plots for the Atrial fibrillation and schizophrenia GWASs. SNPs from LAVA regions that showed significant (FDR-corrected) local genetic correlations are shown in red (correlation was negative) and green (correlation was positive).

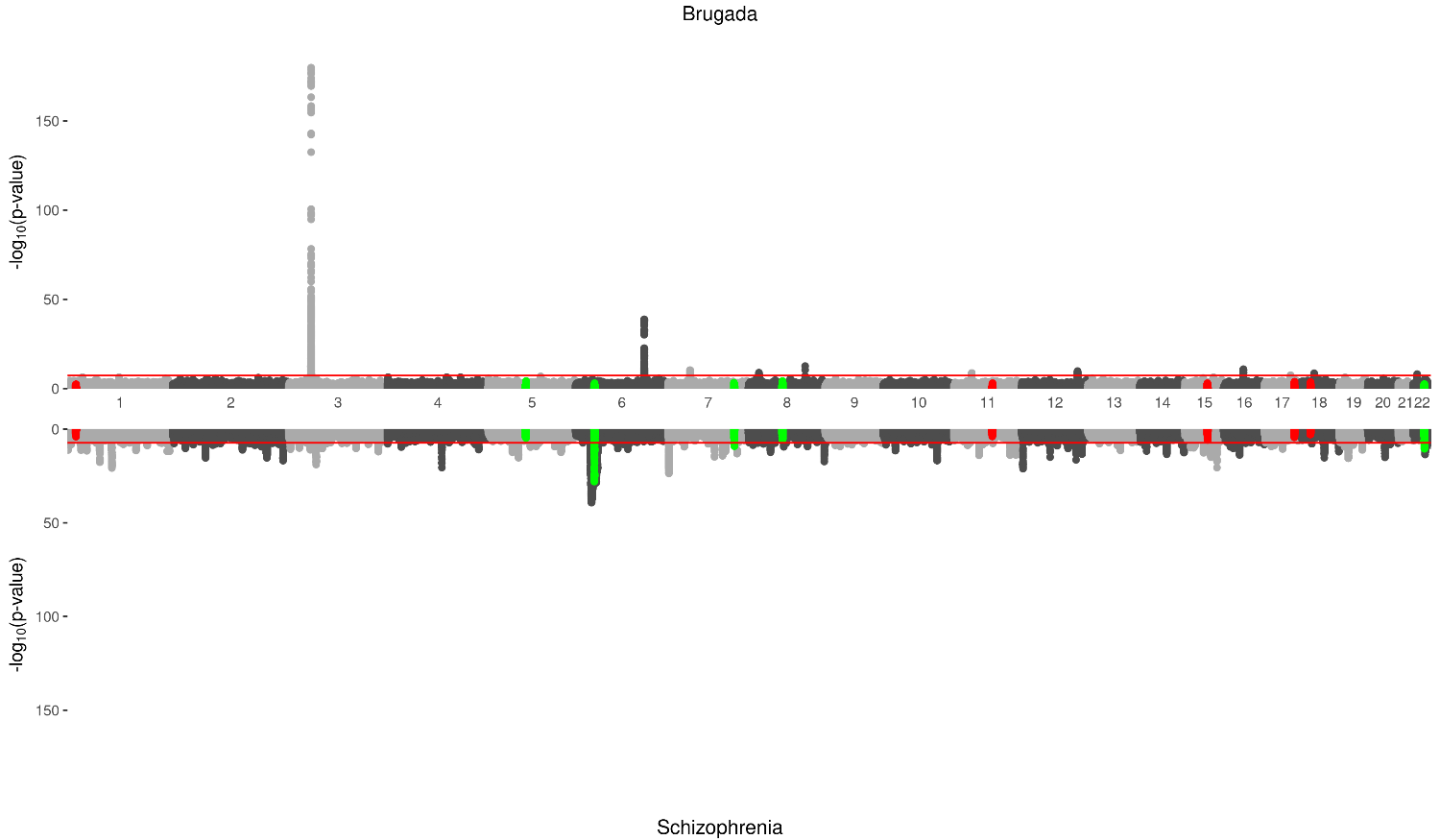

**Figure S3.** Miami plots for the Brugada syndrome and schizophrenia GWASs. SNPs from LAVA regions that showed significant (FDR-corrected) local genetic correlations are shown in red (correlation was negative) and green (correlation was positive).

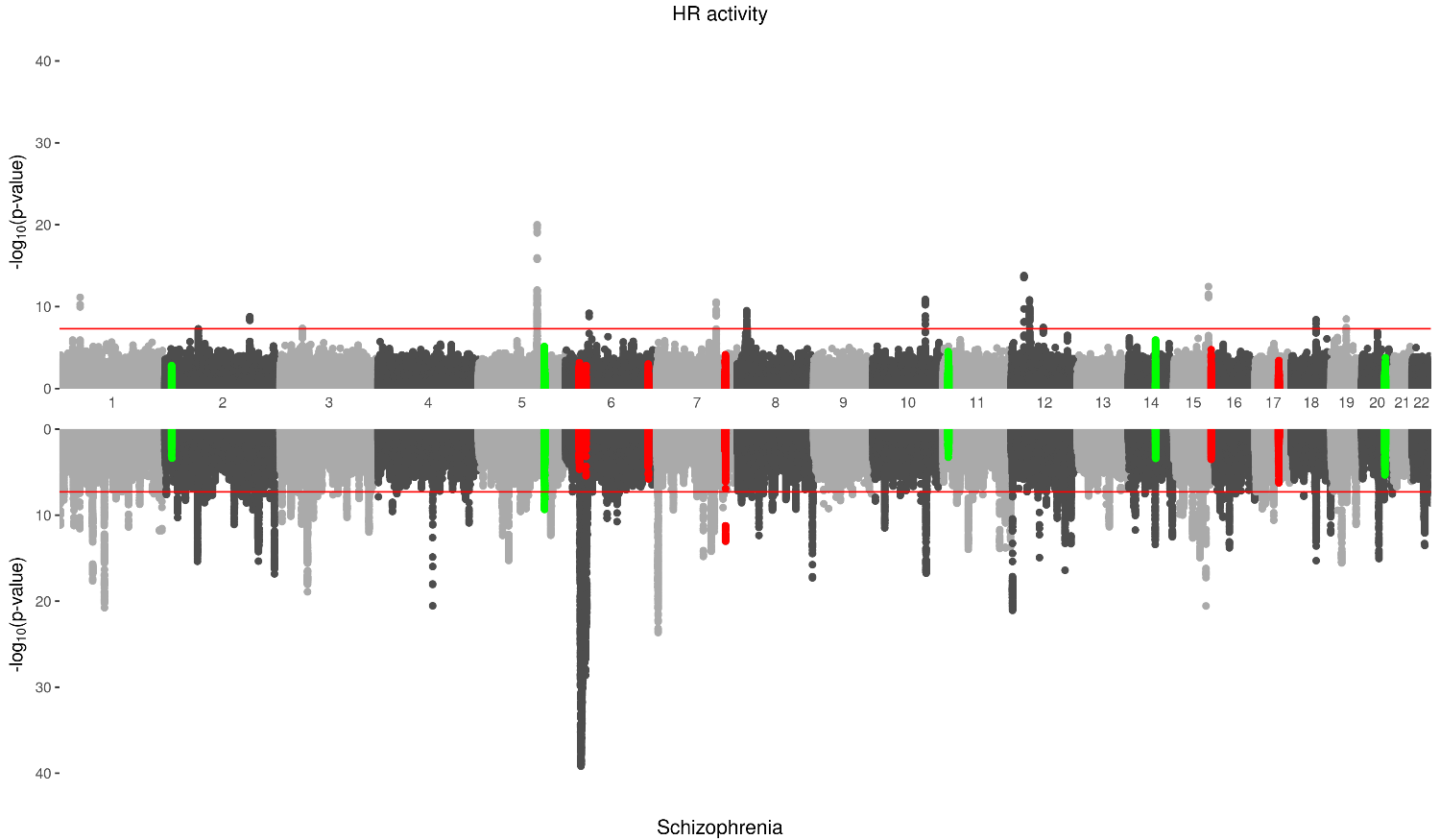

**Figure S4.** Miami plots for the heart rate during activity and schizophrenia GWASs. SNPs from LAVA regions that showed significant (FDR-corrected) local genetic correlations are shown in red (correlation was negative) and green (correlation was positive).

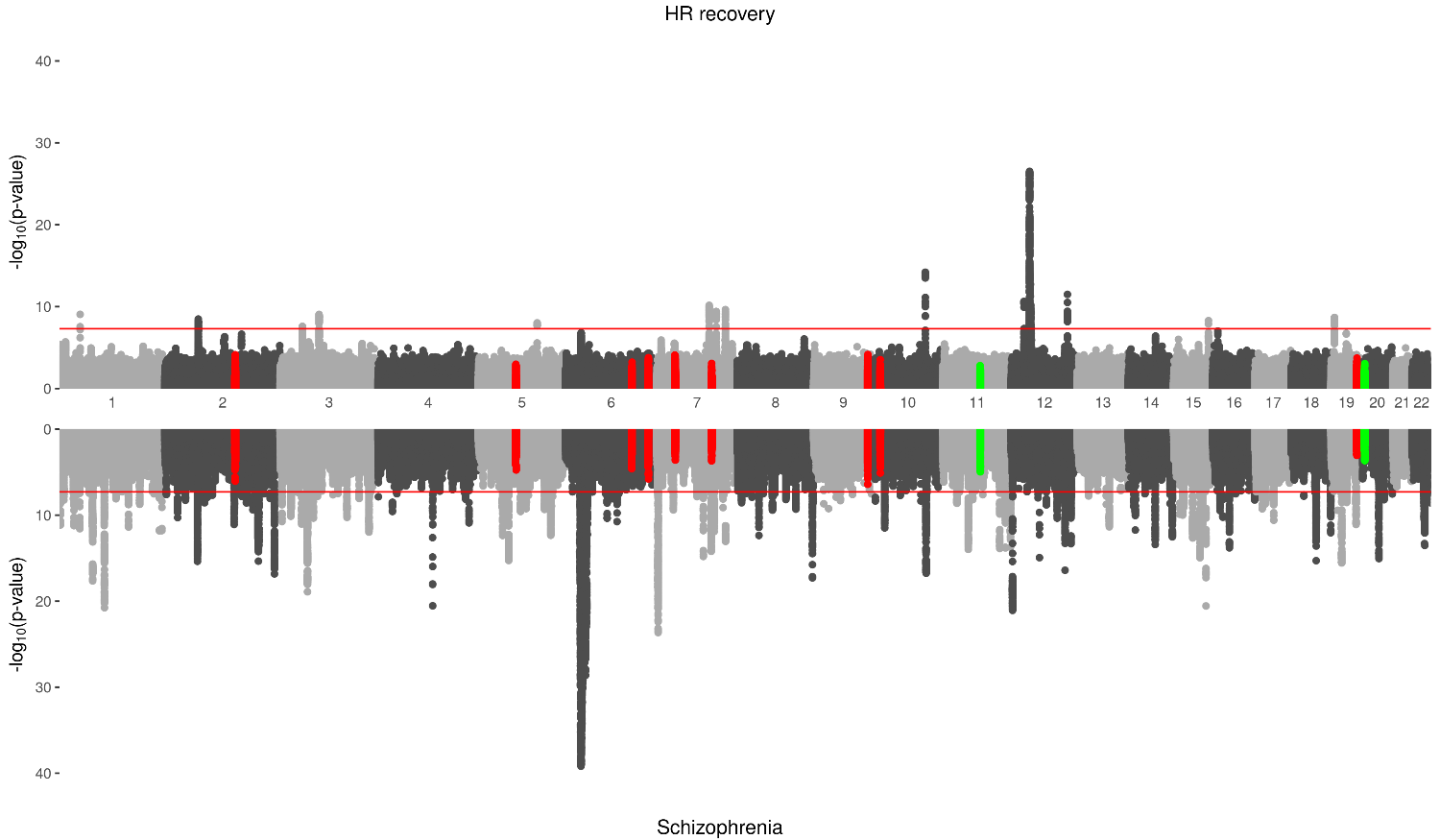

**Figure S5.** Miami plots for the heart rate during recovery and schizophrenia GWASs. SNPs from LAVA regions that showed significant (FDR-corrected) local genetic correlations are shown in red (correlation was negative) and green (correlation was positive).

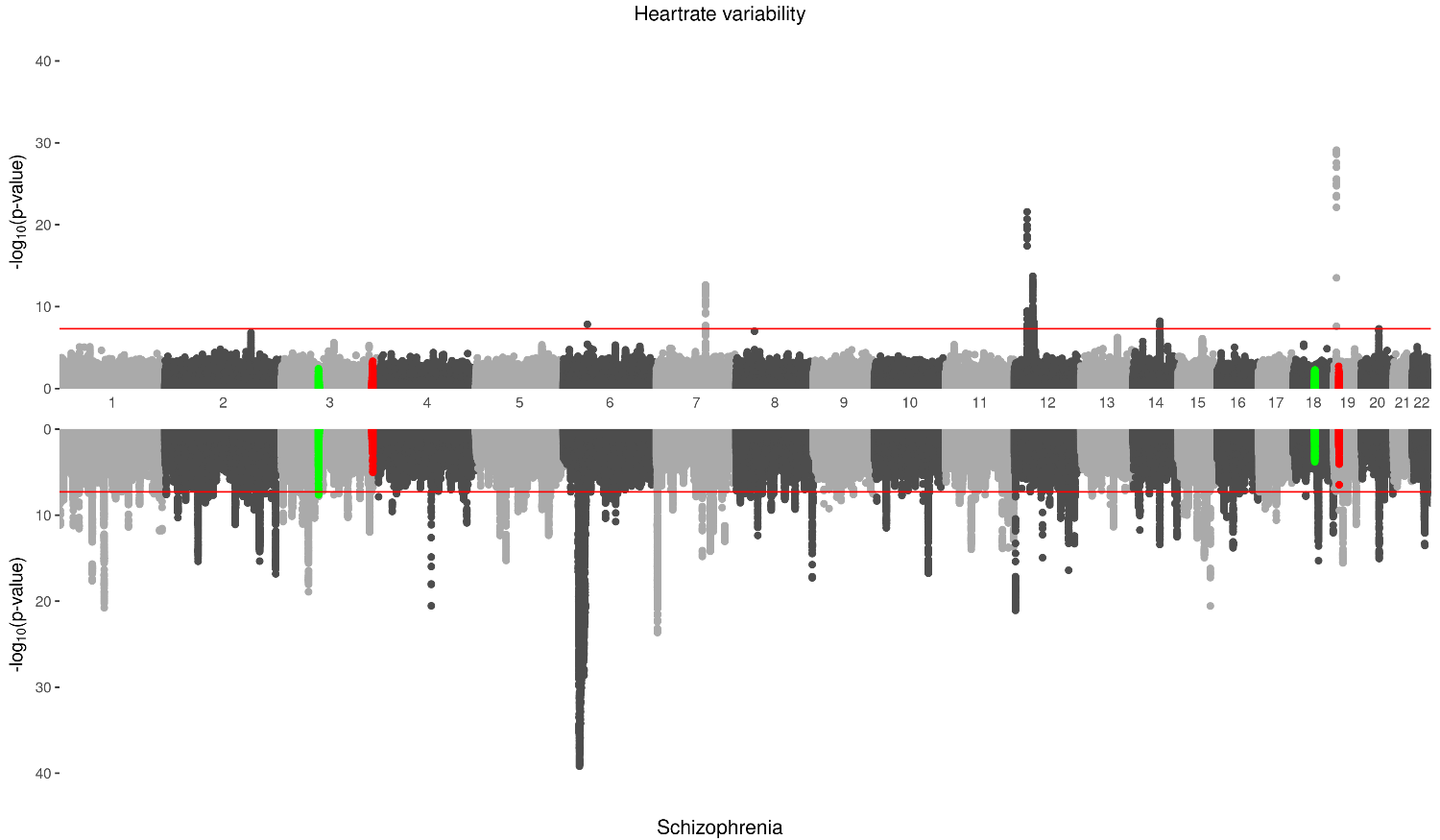

**Figure S6.** Miami plots for the heart rate variability and schizophrenia GWASs. SNPs from LAVA regions that showed significant (FDR-corrected) local genetic correlations are shown in red (correlation was negative) and green (correlation was positive).

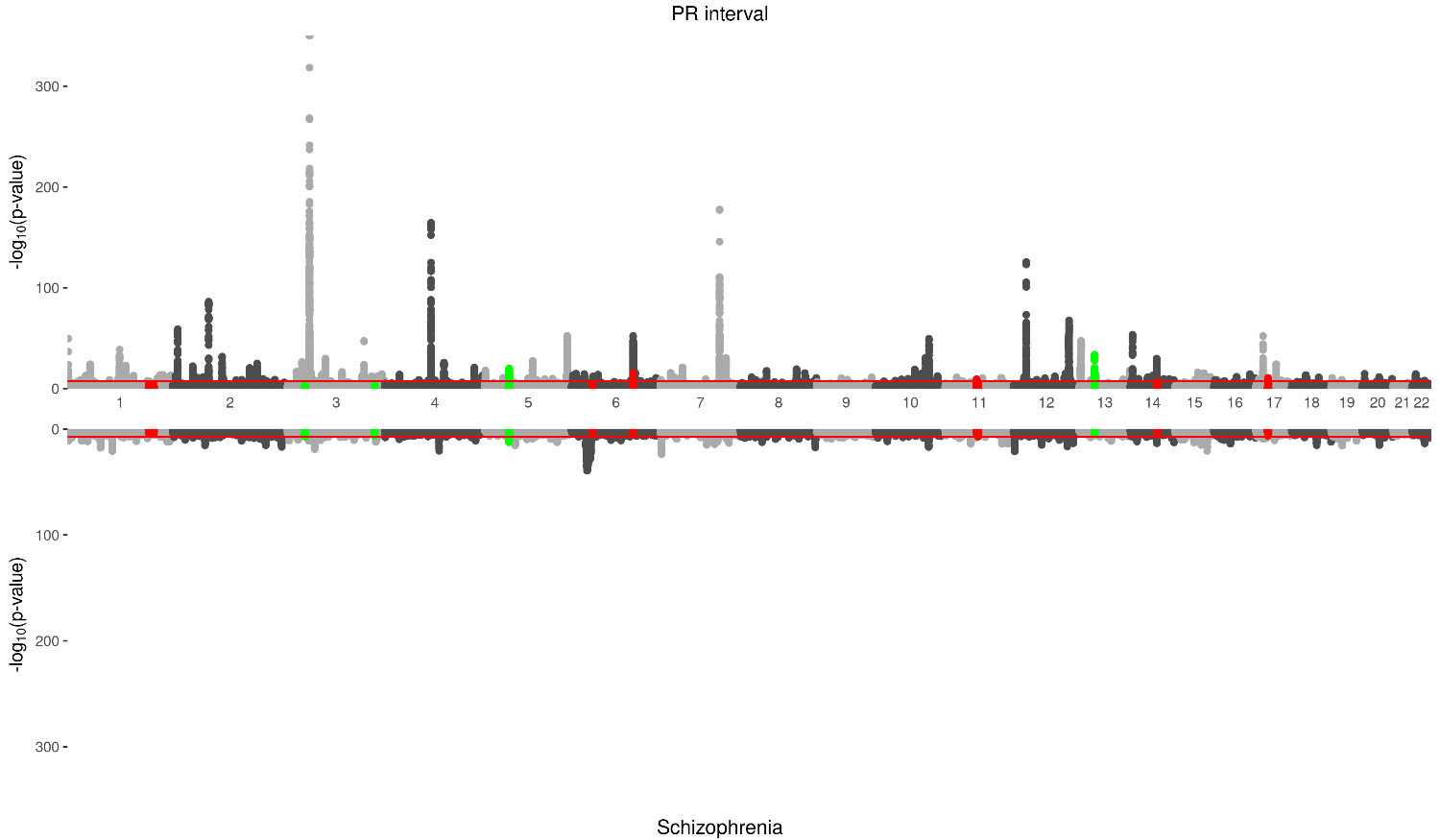

**Figure S7.** Miami plots for the PR interval and schizophrenia GWASs. SNPs from LAVA regions that showed significant (FDR-corrected) local genetic correlations are shown in red (correlation was negative) and green (correlation was positive).

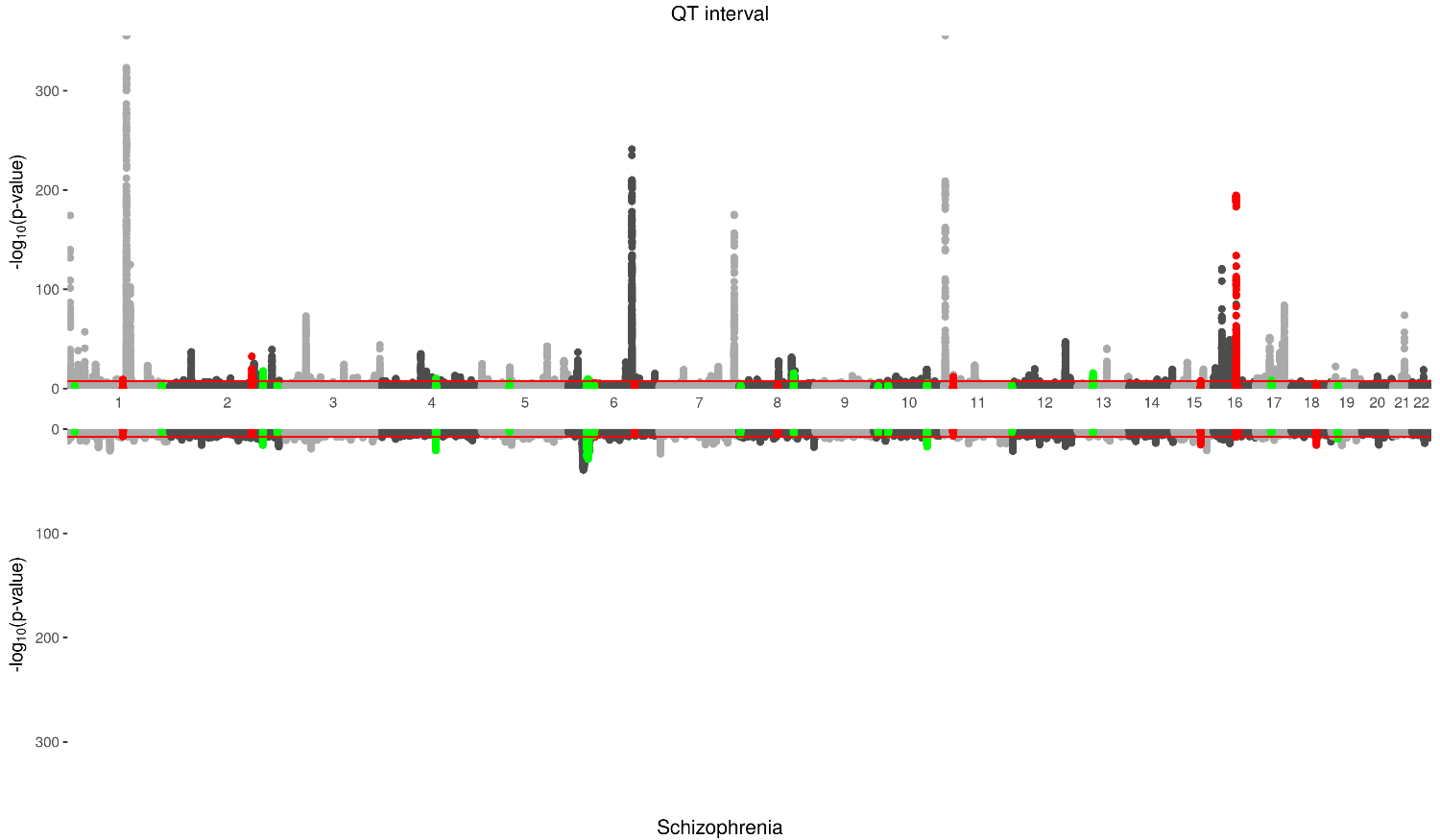

**Figure S8.** Miami plots for the QT interval and schizophrenia GWASs. SNPs from LAVA regions that showed significant (FDR-corrected) local genetic correlations are shown in red (correlation was negative) and green (correlation was positive).

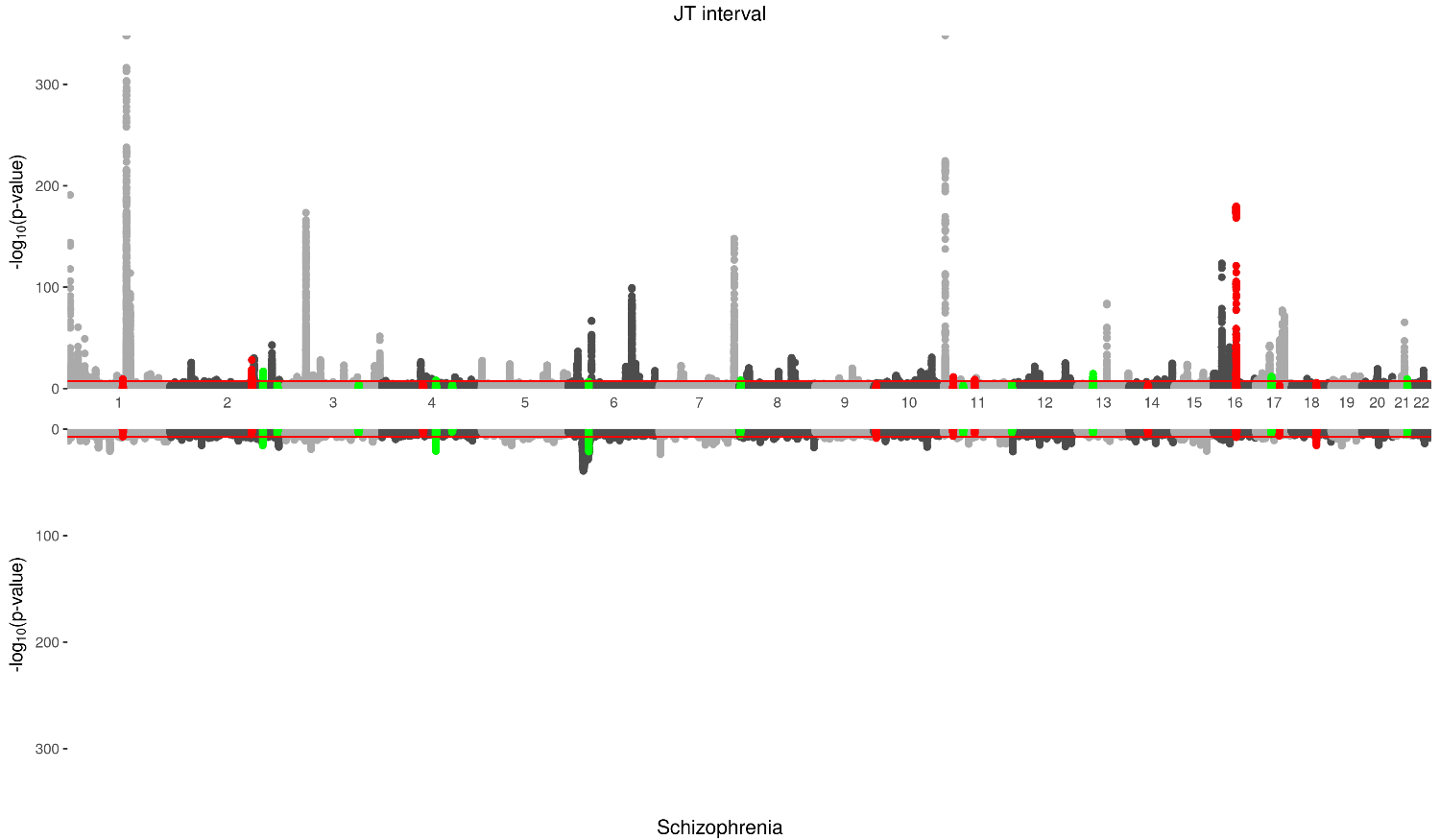

**Figure S9.** Miami plots for the JT interval and schizophrenia GWASs. SNPs from LAVA regions that showed significant (FDR-corrected) local genetic correlations are shown in red (correlation was negative) and green (correlation was positive).

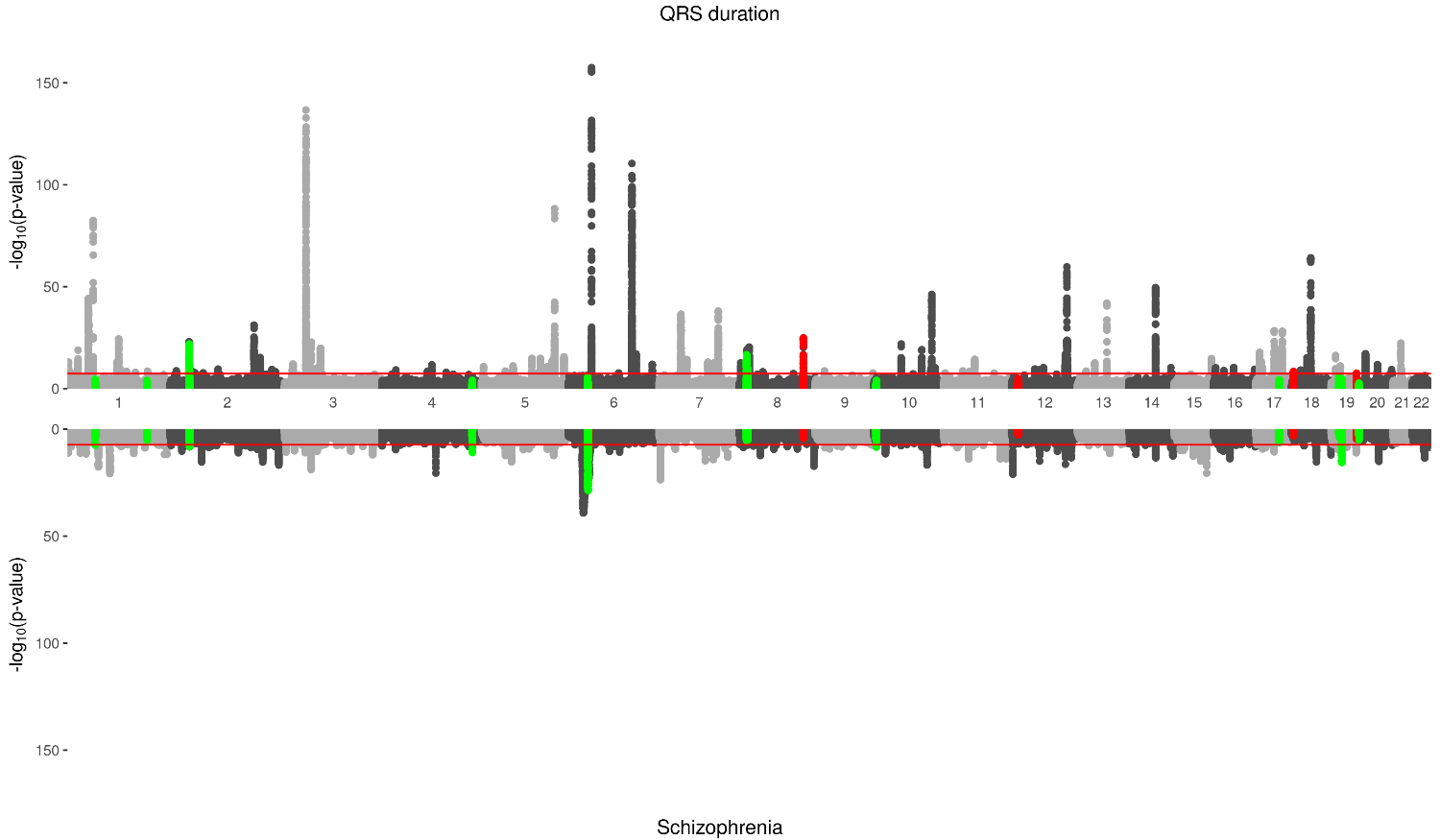

**Figure S10.** Miami plots for the QRS interval and schizophrenia GWASs. SNPs from LAVA regions that showed significant (FDR-corrected) local genetic correlations are shown in red (correlation was negative) and green (correlation was positive).

**Table S1**. Global and MAF-stratified genetic correlations of schizophrenia with all cardiac traits

| **Cardiac traits** | **Global** | | ***MAF 0·05-0·11*** | | **MAF 0·11-0·22** | | **MAF 0·22-0·36** | | **MAF 0·36-0·50** | |
| --- | --- | --- | --- | --- | --- | --- | --- | --- | --- | --- |
|  | **r_g_ (95% CIs)** | ***p-value*** | **r_G_ (95% CIs)** | ***p-value*** | **r_G_ (95% CIs)** | ***p-value*** | **r_G_ (95% CIs)** | ***p-value*** | **r_G_ (95% CIs)** | ***p-value*** |
| AF | -0·034 (-0·075 to -0·007) | 0·106 | -0·073 (-0·165 to 0·020) | 0·125 | -0·017 (-0·085 to 0·051) | 0·621 | -0·027 (-0·087 to 0·034) | 0·388 | -0·021 (-0·085 to 0·043) | 0·523 |
| Brugada | 0·137 (0·061 to 0·213) | 4E-04 | 0·153 (-0·099 to 0·404) | 0·234 | 0·217 (0·048 to 0·385) | 0·012 | 0·163 (0·052 to 0·273) | 0·004 | 0·085 (-0·018 to 0·188) | 0·105 |
| HR act | 0·012 (-0·052 to 0·076) | 0·718 | -0·007 (-0·159 to 0·145) | 0·926 | -0·017 (-0·114 to 0·080) | 0·735 | -0·007 (-0·101 to 0·088) | 0·891 | 0·026 (-0·054 to 0·107) | 0·523 |
| HR rec | -0·072 (-0·142 to -0·002) | 0·044 | -0·128 (-0·299 to0·044) | 0·144 | -0·140 (-0·260 to-0·020) | 0·022 | -0·070 (-0·170 to 0·030) | 0·167 | -0·055 (-0·142 to 0·033) | 0·218 |
| HR var | -0·006 (-0·098 to0·085) | 0·895 | 0·148 (-0·068 to 0·364) | 0·178 | -0·035 (-0·177 to 0·107) | 0·628 | -0·062 (-0·197 to 0·073) | 0·367 | -0·027 (-0·125 to 0·071) | 0·588 |
| PR | -0·023 (-0·061 to 0·015) | 0·242 | -0·036 (-0·120 to 0·048 ) | 0·399 | -0·006 (-0·068 to 0·056) | 0·847 | -0·002 (-0·053 to0·049) | 0·931 | -0·050 (-0·115 to 0·015) | 0·130 |
| QT | 0·019 (-0·020 to 0·058) | 0·343 | -0·031 (-0·109 to 0·048) | 0·442 | 0·026 (-0·043 to 0·095) | 0·463 | 0·041 (-0·015 to 0·098) | 0·153 | 0·027 (-0·026 to 0·081) | 0·316 |
| JT | 0·010 (-0·029 to 0·048 ) | 0·618 | -0·020 (-0·088 to 0·049) | 0·573 | 0·001 (-0·073 to 0·075) | 0·978 | 0·014 (-0·040 to 0·067) | 0·618 | 0·031 (-0·025 to 0·087) | 0·279 |
| QRS | 0·021 (-0·024 to 0·067) | 0·360 | 0·009 (-0·118 to 0·136) | 0·894 | 0·041 (-0·027 to 0·109) | 0·233 | 0·043 (-0·019 to 0·104) | 0·174 | 0·002 (-0·063 to 0·068) | 0·949 |

*AF=Atrial Fibrillation, HR = Heart Rate, MAF=minor allele frequency, r_G_=genetic correlation*

**Table S2**. List of regions showing local correlation (FDR-corrected)

| **Region number** | **Chromosome** | **Start location** | **Stop location** | **n SNPs** | **Cardiac trait** | **r_g_** | ***p*-value** | ***p*-value FDR** |
| --- | --- | --- | --- | --- | --- | --- | --- | --- |
| 379 | 2 | 199654970 | 201324132 | 2558 | AF | 0.313 | 0.000 | 0.000 |
| 849 | 5 | 87943483 | 89584466 | 2073 | AF | 0.238 | 0.001 | 0.046 |
| 1581 | 10 | 104206838 | 106142283 | 3573 | AF | 0.152 | 0.000 | 0.000 |
| 2150 | 16 | 72089512 | 73140781 | 1760 | AF | 0.205 | 0.000 | 0.002 |
| 20 | 1 | 18427821 | 19238924 | 2377 | Brugada | 0.676 | 0.000 | 0.004 |
| 840 | 5 | 77290256 | 79005158 | 3231 | Brugada | 0.303 | 0.000 | 0.025 |
| 957 | 6 | 30715007 | 31106493 | 2359 | Brugada | 0.328 | 0.000 | 0.025 |
| 1209 | 7 | 130418705 | 131856481 | 3251 | Brugada | 0.282 | 0.001 | 0.030 |
| 1294 | 8 | 57436723 | 58675117 | 3348 | Brugada | 0.323 | 0.001 | 0.042 |
| 1686 | 11 | 79802287 | 80726012 | 2292 | Brugada | 0.340 | 0.002 | 0.044 |
| 2072 | 15 | 69089816 | 70767983 | 3252 | Brugada | 0.309 | 0.001 | 0.026 |
| 2228 | 17 | 71466954 | 72383472 | 2383 | Brugada | 0.207 | 0.000 | 0.025 |
| 2258 | 18 | 24088164 | 25305260 | 2670 | Brugada | 0.318 | 0.002 | 0.044 |
| 2483 | 22 | 38718590 | 40378783 | 2744 | Brugada | 0.200 | 0.001 | 0.042 |
| 216 | 2 | 11230350 | 11995765 | 1982 | HR act | 0.378 | 0.003 | 0.041 |
| 891 | 5 | 136949854 | 139408116 | 3133 | HR act | 0.270 | 0.001 | 0.016 |
| 948 | 6 | 23939307 | 24950379 | 2910 | HR act | 0.263 | 0.001 | 0.016 |
| 963 | 6 | 32454578 | 32539567 | 35 | HR act | 0.670 | 0.000 | 0.002 |
| 1080 | 6 | 156762741 | 158220143 | 2971 | HR act | 0.465 | 0.000 | 0.007 |
| 1213 | 7 | 136113468 | 137657459 | 2911 | HR act | 0.317 | 0.000 | 0.009 |
| 1620 | 11 | 7915072 | 9080939 | 3095 | HR act | 0.687 | 0.000 | 0.007 |
| 2002 | 14 | 72665320 | 73373373 | 1880 | HR act | 0.249 | 0.002 | 0.040 |
| 2099 | 15 | 99925973 | 100742117 | 2538 | HR act | 0.471 | 0.001 | 0.013 |
| 2216 | 17 | 54950108 | 56245227 | 2868 | HR act | 0.496 | 0.000 | 0.009 |
| 2413 | 20 | 49236419 | 50653620 | 3818 | HR act | 0.384 | 0.000 | 0.007 |
| 337 | 2 | 147252197 | 149568875 | 3739 | HR rec | 0.315 | 0.000 | 0.013 |
| 840 | 5 | 77290256 | 79005158 | 3333 | HR rec | 0.376 | 0.001 | 0.013 |
| 1052 | 6 | 122929291 | 123854857 | 2184 | HR rec | 0.371 | 0.002 | 0.020 |
| 1080 | 6 | 156762741 | 158220143 | 2971 | HR rec | 1.000 | 0.000 | 0.000 |
| 1126 | 7 | 27351287 | 28890886 | 3541 | HR rec | 0.430 | 0.001 | 0.019 |
| 1191 | 7 | 105312564 | 106399739 | 2733 | HR rec | 0.327 | 0.004 | 0.043 |
| 1474 | 9 | 130902728 | 132193798 | 2025 | HR rec | 0.256 | 0.002 | 0.020 |
| 1498 | 10 | 10969482 | 11856924 | 2327 | HR rec | 0.312 | 0.001 | 0.019 |
| 1682 | 11 | 75445254 | 76518906 | 2238 | HR rec | 0.570 | 0.000 | 0.013 |
| 2358 | 19 | 51259179 | 51903804 | 1876 | HR rec | 0.256 | 0.005 | 0.046 |
| 2374 | 20 | 4591848 | 5379450 | 2393 | HR rec | 0.409 | 0.001 | 0.019 |
| 112 | 1 | 153410810 | 154685545 | 2004 | JT | 0.833 | 0.000 | 0.000 |
| 359 | 2 | 174118390 | 174927563 | 1971 | JT | 0.153 | 0.002 | 0.025 |
| 379 | 2 | 199654970 | 201324132 | 2557 | JT | 0.375 | 0.000 | 0.000 |
| 407 | 2 | 230541093 | 231847386 | 3217 | JT | 0.709 | 0.000 | 0.002 |
| 554 | 3 | 151904022 | 153406718 | 3237 | JT | 0.418 | 0.001 | 0.015 |
| 670 | 4 | 76497359 | 78045637 | 3781 | JT | 0.242 | 0.004 | 0.046 |
| 692 | 4 | 102544804 | 104384534 | 3450 | JT | 0.191 | 0.002 | 0.028 |
| 724 | 4 | 135809543 | 137400031 | 3574 | JT | 0.409 | 0.001 | 0.022 |
| 964 | 6 | 32539568 | 32586784 | 495 | JT | 0.489 | 0.000 | 0.000 |
| 965 | 6 | 32586785 | 32629239 | 678 | JT | 0.493 | 0.000 | 0.000 |
| 1235 | 8 | 1628040 | 2070902 | 1519 | JT | 0.379 | 0.001 | 0.015 |
| 1489 | 10 | 3225337 | 3880322 | 1843 | JT | 0.229 | 0.002 | 0.029 |
| 1628 | 11 | 16383387 | 17583948 | 2383 | JT | 0.444 | 0.000 | 0.001 |
| 1648 | 11 | 35500368 | 36331189 | 1934 | JT | 0.361 | 0.003 | 0.039 |
| 1671 | 11 | 60515106 | 61717117 | 1975 | JT | 0.207 | 0.003 | 0.036 |
| 1740 | 12 | 60317 | 1078397 | 1366 | JT | 0.213 | 0.004 | 0.045 |
| 1889 | 13 | 46493237 | 47433528 | 2042 | JT | 0.390 | 0.000 | 0.010 |
| 1988 | 14 | 56206431 | 57460781 | 3319 | JT | 0.164 | 0.003 | 0.033 |
| 2139 | 16 | 58508979 | 59885499 | 3527 | JT | 0.302 | 0.000 | 0.000 |
| 2203 | 17 | 37361179 | 38880481 | 2387 | JT | 0.286 | 0.000 | 0.003 |
| 2216 | 17 | 54950108 | 56245227 | 2880 | JT | 0.366 | 0.002 | 0.025 |
| 2281 | 18 | 52512524 | 53762996 | 2161 | JT | 0.223 | 0.000 | 0.010 |
| 2454 | 21 | 40480832 | 41344742 | 2645 | JT | 0.341 | 0.001 | 0.015 |
| 156 | 1 | 202583885 | 204092537 | 3276 | PR | 0.374 | 0.001 | 0.027 |
| 163 | 1 | 212347583 | 213958292 | 3631 | PR | 0.619 | 0.000 | 0.012 |
| 449 | 3 | 28649811 | 29877022 | 3633 | PR | 0.267 | 0.002 | 0.045 |
| 577 | 3 | 176931164 | 178110322 | 2503 | PR | 0.255 | 0.002 | 0.046 |
| 814 | 5 | 44049879 | 45305566 | 1873 | PR | 0.587 | 0.000 | 0.002 |
| 815 | 5 | 45305567 | 46388848 | 1784 | PR | 0.457 | 0.000 | 0.011 |
| 971 | 6 | 34979271 | 36346353 | 2611 | PR | 0.275 | 0.001 | 0.025 |
| 1046 | 6 | 117674077 | 118529071 | 1882 | PR | 0.253 | 0.001 | 0.025 |
| 1671 | 11 | 60515106 | 61717117 | 1963 | PR | 0.287 | 0.001 | 0.025 |
| 1674 | 11 | 64594823 | 66782661 | 3031 | PR | 0.289 | 0.002 | 0.043 |
| 1889 | 13 | 46493237 | 47433528 | 2027 | PR | 0.433 | 0.000 | 0.001 |
| 2002 | 14 | 72665320 | 73373373 | 1884 | PR | 0.325 | 0.001 | 0.026 |
| 2196 | 17 | 27344402 | 29783141 | 3036 | PR | 0.382 | 0.002 | 0.045 |
| 56 | 1 | 65894185 | 66778015 | 1739 | QRS | 0.265 | 0.002 | 0.031 |
| 157 | 1 | 204092538 | 205009623 | 2072 | QRS | 0.321 | 0.001 | 0.018 |
| 241 | 2 | 36867390 | 37603360 | 1988 | QRS | 0.644 | 0.000 | 0.001 |
| 758 | 4 | 175959698 | 177129678 | 2637 | QRS | 0.189 | 0.002 | 0.026 |
| 961 | 6 | 31427210 | 32208901 | 2274 | QRS | 0.217 | 0.000 | 0.012 |
| 964 | 6 | 32539568 | 32586784 | 495 | QRS | 0.197 | 0.000 | 0.008 |
| 1246 | 8 | 8064601 | 8589770 | 1573 | QRS | 0.209 | 0.002 | 0.031 |
| 1248 | 8 | 9167796 | 9835863 | 1795 | QRS | 0.338 | 0.004 | 0.050 |
| 1350 | 8 | 124001020 | 125453322 | 3282 | QRS | 0.397 | 0.000 | 0.000 |
| 1489 | 10 | 3225337 | 3880322 | 1843 | QRS | 0.242 | 0.002 | 0.026 |
| 1753 | 12 | 12721875 | 13559527 | 1700 | QRS | 0.291 | 0.001 | 0.018 |
| 2216 | 17 | 54950108 | 56245227 | 2880 | QRS | 0.513 | 0.000 | 0.008 |
| 2241 | 18 | 2839843 | 3722828 | 2355 | QRS | 0.254 | 0.001 | 0.015 |
| 2321 | 19 | 13968320 | 14891100 | 1988 | QRS | 0.488 | 0.000 | 0.004 |
| 2327 | 19 | 18504868 | 19749166 | 2171 | QRS | 0.213 | 0.001 | 0.020 |
| 2357 | 19 | 50451926 | 51259178 | 1824 | QRS | 0.262 | 0.000 | 0.009 |
| 2362 | 19 | 54042241 | 54602370 | 1760 | QRS | 0.423 | 0.001 | 0.018 |
| 18 | 1 | 16732169 | 17557746 | 1055 | QT | 0.407 | 0.003 | 0.031 |
| 112 | 1 | 153410810 | 154685545 | 2004 | QT | 0.692 | 0.000 | 0.001 |
| 184 | 1 | 234365797 | 235097113 | 2056 | QT | 0.247 | 0.006 | 0.045 |
| 359 | 2 | 174118390 | 174927563 | 1971 | QT | 0.174 | 0.001 | 0.009 |
| 379 | 2 | 199654970 | 201324132 | 2557 | QT | 0.395 | 0.000 | 0.000 |
| 407 | 2 | 230541093 | 231847386 | 3217 | QT | 0.605 | 0.000 | 0.002 |
| 692 | 4 | 102544804 | 104384534 | 3450 | QT | 0.330 | 0.000 | 0.002 |
| 823 | 5 | 55968967 | 56896890 | 2065 | QT | 0.311 | 0.003 | 0.029 |
| 958 | 6 | 31106494 | 31250556 | 1728 | QT | 0.332 | 0.001 | 0.008 |
| 959 | 6 | 31250557 | 31320268 | 1337 | QT | 0.735 | 0.000 | 0.000 |
| 960 | 6 | 31320269 | 31427209 | 1554 | QT | 0.375 | 0.000 | 0.001 |
| 961 | 6 | 31427210 | 32208901 | 2274 | QT | 0.261 | 0.000 | 0.001 |
| 964 | 6 | 32539568 | 32586784 | 495 | QT | 0.421 | 0.000 | 0.000 |
| 965 | 6 | 32586785 | 32629239 | 678 | QT | 0.471 | 0.000 | 0.000 |
| 966 | 6 | 32629240 | 32682213 | 776 | QT | 0.297 | 0.000 | 0.001 |
| 976 | 6 | 40778402 | 42103738 | 3117 | QT | 0.230 | 0.002 | 0.022 |
| 977 | 6 | 42103739 | 43770626 | 3153 | QT | 0.246 | 0.001 | 0.010 |
| 1052 | 6 | 122929291 | 123854857 | 2185 | QT | 0.210 | 0.003 | 0.031 |
| 1235 | 8 | 1628040 | 2070902 | 1519 | QT | 0.937 | 0.000 | 0.000 |
| 1304 | 8 | 68683824 | 70004665 | 3177 | QT | 0.664 | 0.000 | 0.001 |
| 1332 | 8 | 103716054 | 104467845 | 1953 | QT | 0.226 | 0.004 | 0.036 |
| 1493 | 10 | 6234719 | 6897037 | 1754 | QT | 0.177 | 0.006 | 0.045 |
| 1511 | 10 | 23365686 | 24489447 | 2150 | QT | 0.470 | 0.002 | 0.021 |
| 1581 | 10 | 104206838 | 106142283 | 3571 | QT | 0.203 | 0.001 | 0.018 |
| 1628 | 11 | 16383387 | 17583948 | 2383 | QT | 0.489 | 0.000 | 0.000 |
| 1740 | 12 | 60317 | 1078397 | 1366 | QT | 0.352 | 0.000 | 0.008 |
| 1889 | 13 | 46493237 | 47433528 | 2042 | QT | 0.311 | 0.001 | 0.011 |
| 2079 | 15 | 78514102 | 79292536 | 1832 | QT | 0.611 | 0.000 | 0.000 |
| 2139 | 16 | 58508979 | 59885499 | 3527 | QT | 0.281 | 0.000 | 0.000 |
| 2141 | 16 | 61092845 | 62607090 | 2690 | QT | 0.327 | 0.002 | 0.022 |
| 2203 | 17 | 37361179 | 38880481 | 2387 | QT | 0.187 | 0.004 | 0.034 |
| 2281 | 18 | 52512524 | 53762996 | 2161 | QT | 0.219 | 0.002 | 0.021 |
| 2319 | 19 | 11681979 | 13213186 | 2745 | QT | 0.221 | 0.005 | 0.045 |
| 483 | 3 | 71223282 | 72334704 | 981 | HRV | 0.456 | 0.000 | 0.001 |
| 584 | 3 | 186602046 | 187939199 | 1305 | HRV | 0.296 | 0.001 | 0.045 |
| 2274 | 18 | 45718493 | 46558306 | 735 | HRV | 0.467 | 0.001 | 0.045 |
| 2318 | 19 | 10028841 | 11681978 | 672 | HRV | 0.540 | 0.000 | 0.001 |

3. **Functional annotation analyses**

**A**

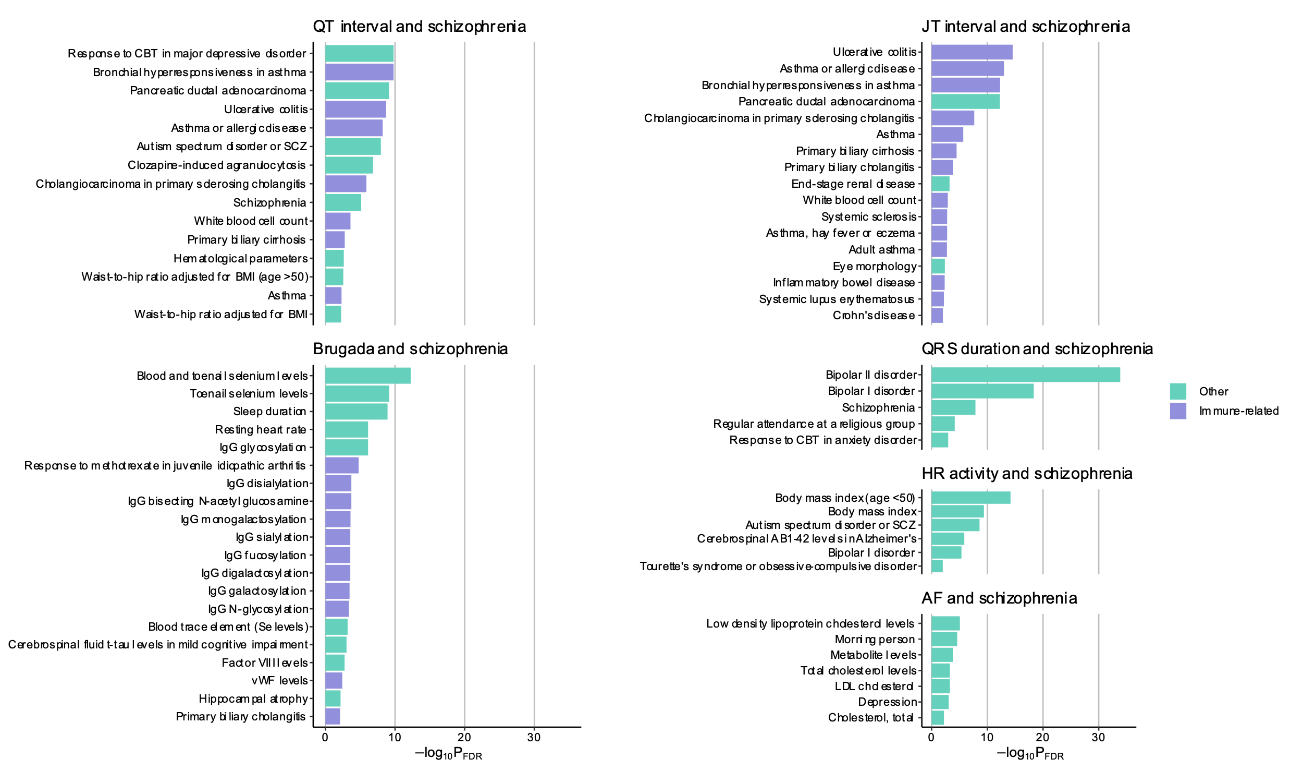

**B**

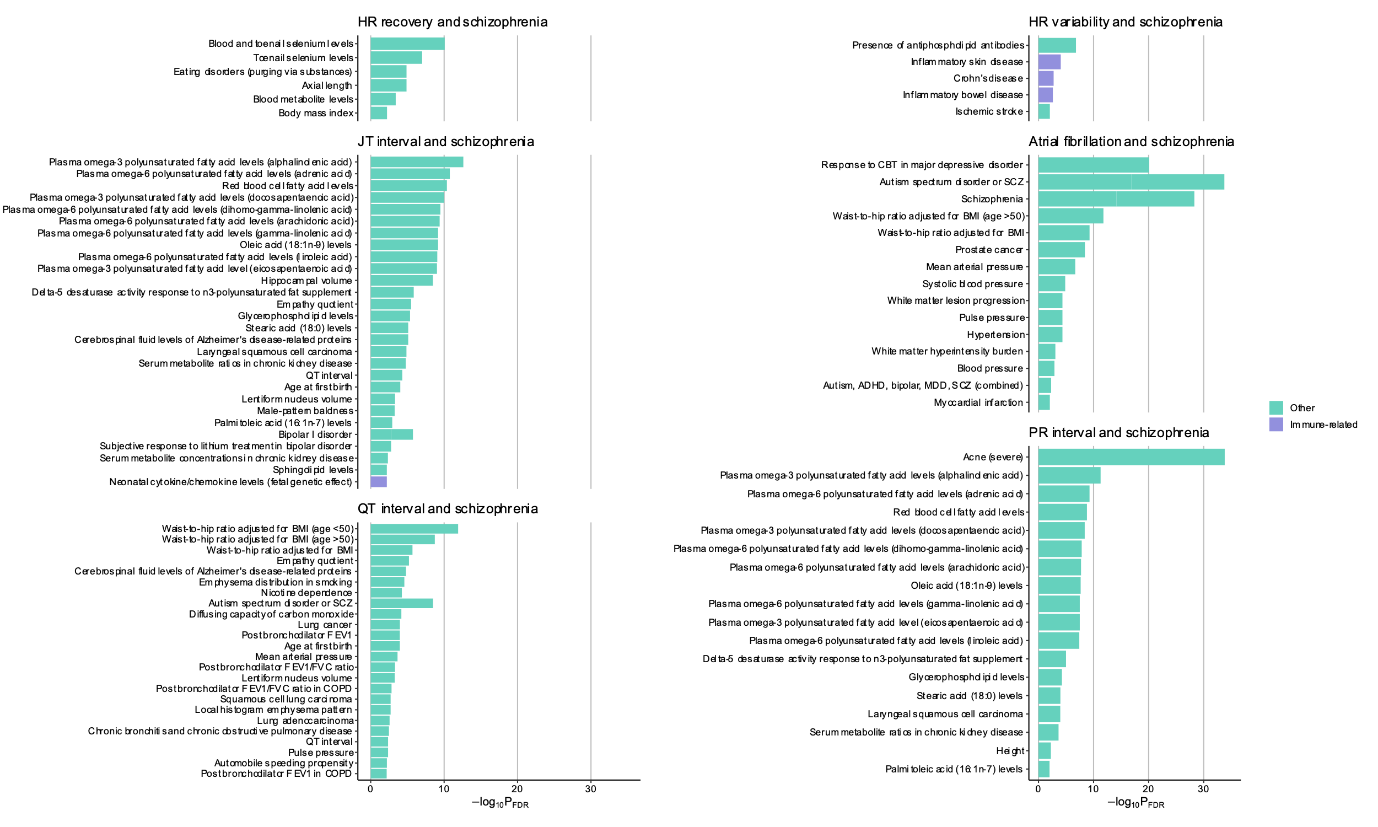

**Figure S11.** Previously reported trait associations from GWAS catalogue for genes in regions with a significant SCZ-cardiac trait correlation (excluding HLA region). **Panel A)** shows results for genes in regions with a positive SCZ-cardiac trait correlation; **panel B)** shows results for genes in regions with a negative correlation.

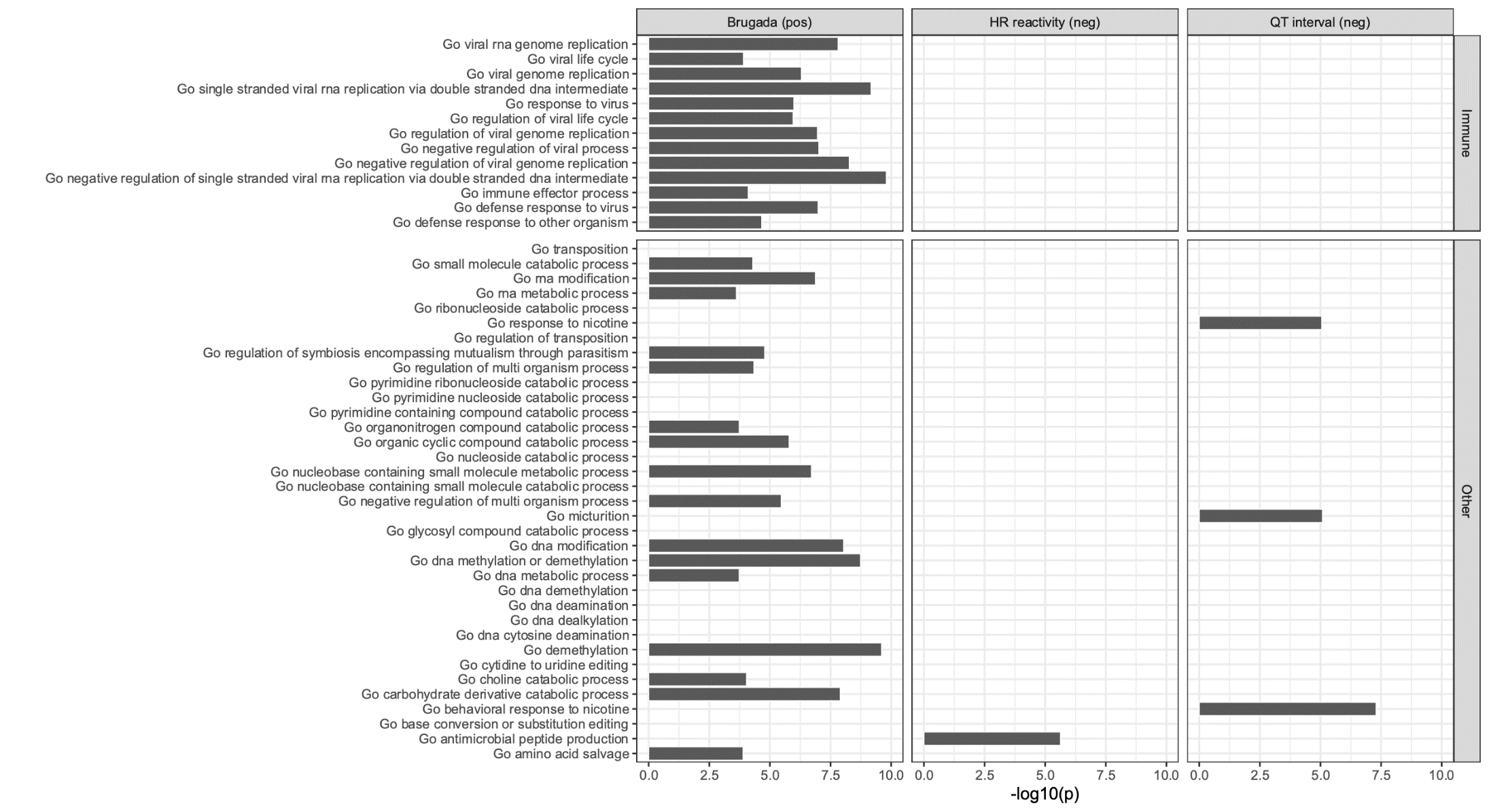

**Figure S12.** Full results of GO biological processes enrichment for genes in regions with a significant SCZ-cardiac trait association (excluding HLA region). Only results for trait pairs with any significant enrichment are plotted. Results for processes associated with immune function or viral response (from different GO branches) are shown at the top.

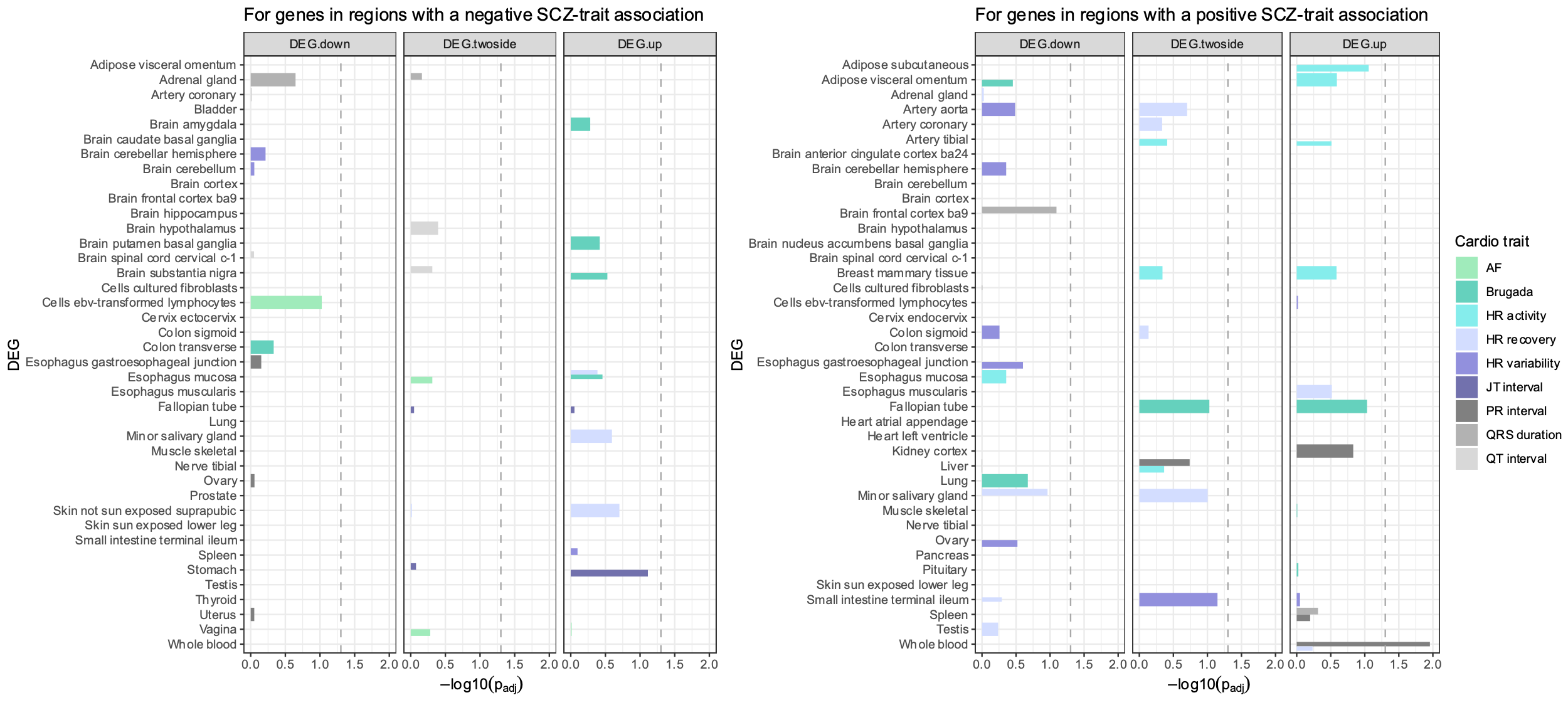

**Figure S13.** Full results for differential tissue expression across the 30 available tissue types of the GTEx project for genes in regions with a significant SCZ-cardiac trait association (excluding HLA region), with the left panel showing results for genes in negatively associated regions, and the right panel for genes in positively associated reg

4. **Mendelian randomization analyses**

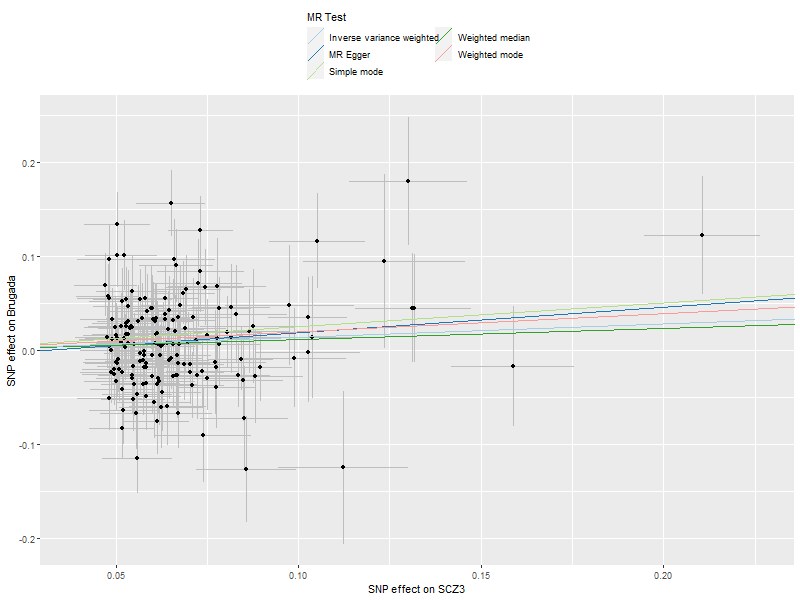

**Figure S14**. Scatter plot for the Mendelian randomization results with liability to schizophrenia as the exposure and Brugada syndrome as the outcome.

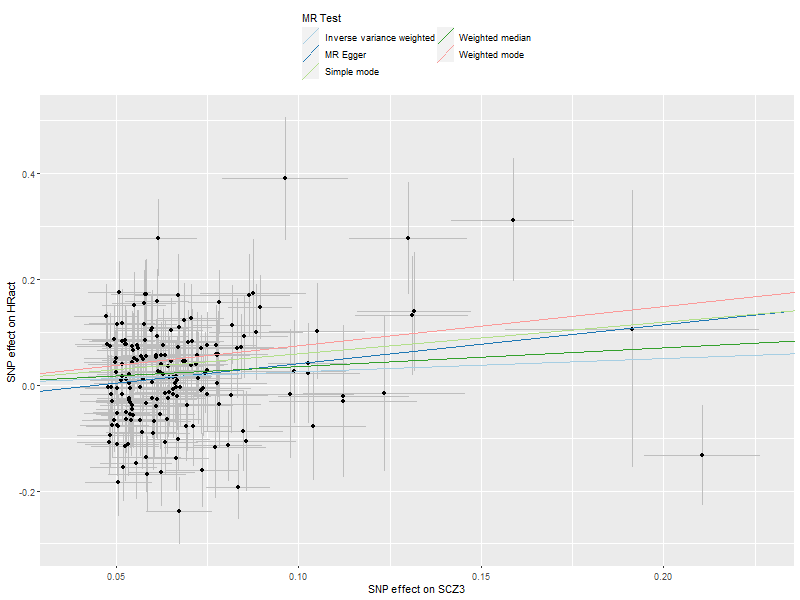

**Figure S15**. Scatter plot for the Mendelian randomization results with liability to schizophrenia as the exposure and heart rate during activity as the outcome.

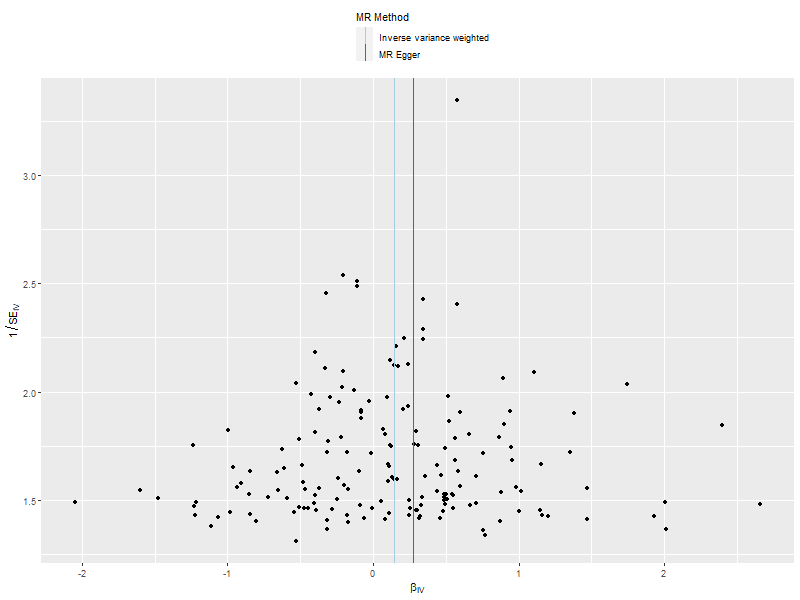

**Figure S16.** Funnel plot for the Mendelian randomization results with liability to schizophrenia as the exposure and Brugada syndrome as the outcome

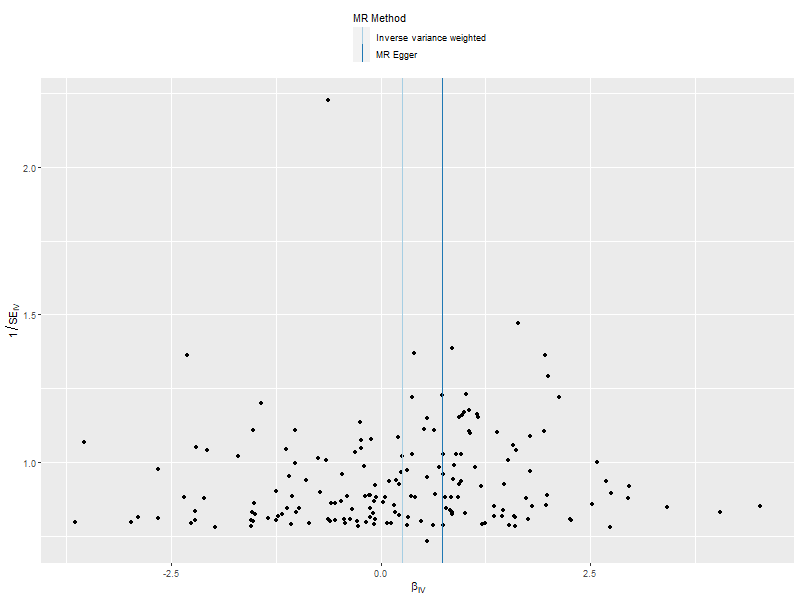

**Figure S17.** Funnel plot for the Mendelian randomization results with liability to schizophrenia as the exposure and heart rate during activity as the outcome.

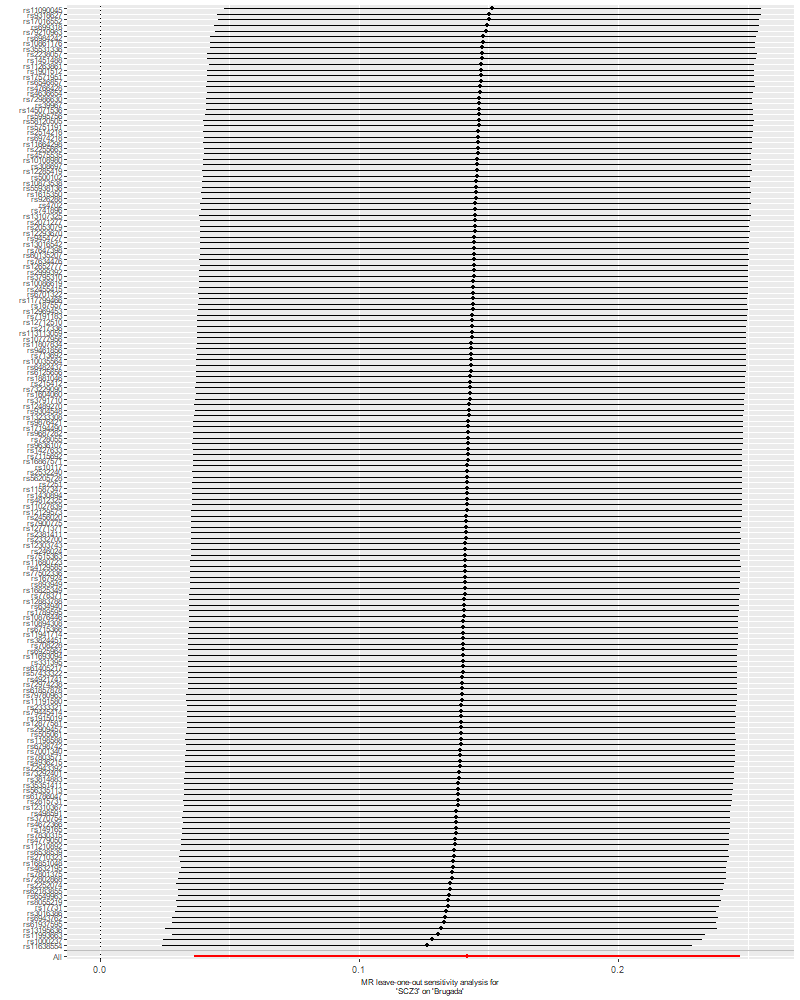

**Figure S18.** Leave-one-out analysis for the Mendelian randomization results with liability to schizophrenia as the exposure and Brugada syndrome as the outcome.

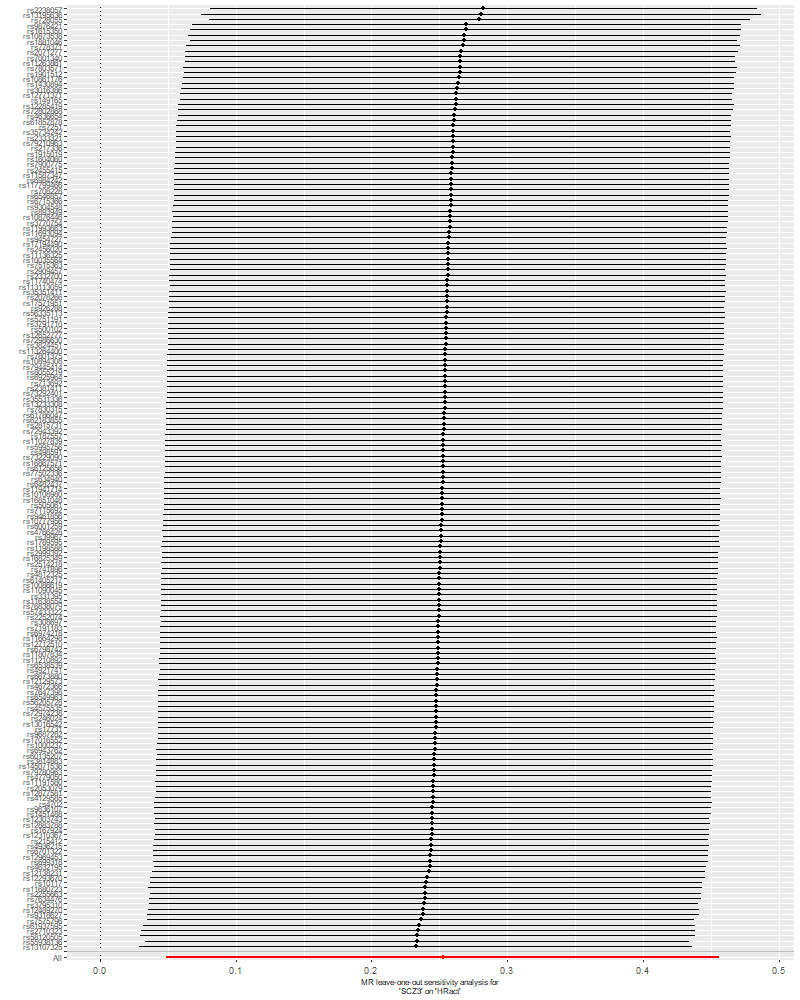

**Figure S19.** Leave-one-out analysis for the Mendelian randomization results with liability to schizophrenia as the exposure and heart rate during activity as the outcome

**Table S3**. Test for heterogeneity with Cochran’s Q statistic for all univariable MR analyses

| **Exposure** | **Outcome** | **Method** | **Q** | **df** | **P value** |
| --- | --- | --- | --- | --- | --- |
| Schizophrenia | AF | IVW | 280.14 | 151 | 8.7E-10 |
|  |  | MR Egger | 279.72 | 150 | 6.9E-10 |
| Schizophrenia | Brugada | IVW | 243.44 | 168 | 1.2E-04 |
|  |  | MR Egger | 242.94 | 167 | 1.3E-04 |
| Schizophrenia | HR activity | IVW | 307.00 | 176 | 3.6E-09 |
|  |  | MR Egger | 304.56 | 175 | 4.7E-09 |
| Schizophrenia | HR recovery | IVW | 238.03 | 176 | 0.001 |
|  |  | MR Egger | 237.61 | 176 | 0.001 |
| Schizophrenia | HR variability | IVW | 118.33 | 92 | 0.034 |
|  |  | MR Egger | 118.31 | 91 | 0.029 |
| Schizophrenia | PR interval | IVW | 378.37 | 172 | 1.5E-17 |
|  |  | MR Egger | 378.31 | 171 | 1.5E-17 |
| Schizophrenia | QT interval | IVW | 699.96 | 173 | 1.7E-64 |
|  |  | MR Egger | 698.29 | 172 | 1.6E-64 |
| Schizophrenia | JT interval | IVW | 596.52 | 175 | 2.1E-47 |
|  |  | MR Egger | 596.52 | 174 | 1.2E-47 |
| Schizophrenia | QRS duration | IVW | 438.61 | 175 | 1.3E-24 |
|  |  | MR Egger | 430.22 | 174 | 1.1E-23 |
| AF | Schizophrenia | IVW | 172.94 | 72 | 2.8E-10 |
|  |  | MR Egger | 170.99 | 71 | 3.2E-10 |
| Brugada | Schizophrenia | IVW | 21.67 | 11 | 0.027 |
|  |  | MR Egger | 19.44 | 10 | 0.035 |
| HR activity | Schizophrenia | IVW | 18.34 | 11 | 0.074 |
|  |  | MR Egger | 18.03 | 10 | 0.055 |
| HR recovery | Schizophrenia | IVW | 24.88 | 11 | 0.009 |
|  |  | MR Egger | 20.96 | 10 | 0.021 |
| HR variability | Schizophrenia | IVW | 11.48 | 8 | 0.176 |
|  |  | MR Egger | 11.25 | 7 | 0.128 |
| PR interval | Schizophrenia | IVW | 552.62 | 253 | 2.1E-24 |
|  |  | MR Egger | 552.62 | 252 | 1.4E-24 |
| QT interval | Schizophrenia | IVW | 496.88 | 151 | 1.8E-38 |
|  |  | MR Egger | 495.96 | 150 | 1.3E-38 |
| JT interval | Schizophrenia | IVW | 309.37 | 135 | 1.0E-15 |
|  |  | MR Egger | 303.89 | 134 | 3.2E-15 |
| QRS duration | Schizophrenia | IVW | 253.28 | 99 | 1.8E-15 |
|  |  | MR Egger | 251.81 | 98 | 1.7E-15 |

| **Exposure** | **Outcome** | **Additional exposure** | **IVW**  **beta (95% Cis), p** |
| --- | --- | --- | --- |
| Schizophrenia | Brugada |  | 0.14 (0.03 to 0.25), 0.009 |
|  | Brugada | HR activity | 0.19 (-0.02 to 0.40), 0.089 |
|  | Brugada | HR recovery | 0.17 (-0.04 to 0.38), 0.123 |
|  | Brugada | HR variability | 0.17 (0.03 to 0.31), 0.015 |
|  | Brugada | PR interval | 0.13 (-0.04 to 0.30), 0.125 |
|  | Brugada | QT interval | 0.15 (0.01 to 0.29), 0.042 |
|  | Brugada | JT interval | 0.16 (0.00 to 0.32), 0.042 |
|  | Brugada | QRS duration | 0.12 (-0.01 to 0.25), 0.076 |

**Table S4.** Multivariable Mendelian randomization

*Note that the Inverse Variance Weighted (IVW) estimate represents the causal effect of liability to schizophrenia on Brugada, before and after adding each of the additional exposures separately.*
